# Inference on the dynamics of the COVID pandemic from observational data

**DOI:** 10.1101/2021.02.01.21250936

**Authors:** Satarupa Bhattacharjee, Shuting Liao, Debashis Paul, Sanjay Chaudhuri

**Affiliations:** Department of Statistics, University of California, Davis; Graduate Group in BioStatistics, University of California, Davis; Department of Statistics and Applied Probability, National University of Singapore

**Keywords:** COVID-19, Stochastic Dynamic Model, Nonparametric Inference, Multiple Compartments

## Abstract

We describe a time dependent stochastic dynamic model in discrete time for the evolution of the COVID-19 pandemic in various states of USA. The proposed multi-compartment model is expressed through a system of difference equations that describe their temporal dynamics. Various compartments in our model is connected to the social distancing measures and diagnostic testing rates. A nonparametric estimation strategy is employed for obtaining estimates of interpretable temporally static and dynamic epidemiological rate parameters. The confidence bands of the parameters are obtained using a residual bootstrap procedure. A key feature of the methodology is its ability to estimate latent compartments such as the trajectory of the number of asymptomatic but infected individuals which are the key vectors of COVID-19 spread. The nature of the disease dynamics is further quantified by the proposed epidemiological markers, which use estimates of such key latent compartments.

## 1. Introduction

The novel coronavirus has been ravaging the world since early 2020. First identified in Wuhan, Hubei Province, China, the epidemic has since spread to every corner of the world. As of January 25, 2021 [1], more than 99 million people have been infected, out of which more than 2.1 million have died of the disease. The World Health Organization declared the situation a pandemic on March 11, 2020. Since then various parts of the world have gone through multiple waves surges in the number of new infections. The pandemic has severely affected the world economy. Repeated lockdowns, travel restrictions, and other measures of containment have severely impacted the economy of many countries, stretched healthcare systems to the extreme, and caused mental health crisis for large chunks of the populations.

The new pathogen (SARS-CoV-2) which causes the disease [2] is mostly unknown in terms of its infectivity and clinical profile. It is well-known that the infection primarily spreads through infected but asymptomatic people. The number of such people remains unknown. The reported number is based on symptomatic or positively tested persons, which grossly underestimates of the true value. Because of the undetermined denominator effect, important epidemiological markers like the death rate, hospitalisation rate etc remains non-determinable from the observed data. Various estimates [3, 4, 5, 6, 7] of these markers have been postulated by many authors. Mathematical modelling and quantification of the epidemiological parameters [8, 9, 10, 11, 12, 13] of the pandemic have been crucial in understanding and interpreting the transmission dynamics from the perspective of public health researchers and policymakers around the globe [14, 15, 16, 17].

A number of popular compartmental epidemiological models e.g. SIR, SEIR, SIRD etc. have been employed to describe the dynamics of COVID-19 [18, 19, 20, 21]. Such models yield estimates of epidemilogical markers such as the basic reproduction number (*R*_0_), and various doubling and case fatality rates that are indicators of the disease growth pattern [22, 23]. Prediction of epidemiological characteristics and transmission patterns in this context have also attracted major attention [24, 25, 26, 27]. Advanced statistical methods have been employed in forecasting the number of cases worldwide [28] or quantifying the effects of prevention mechanisms like social distancing [29, 30, 31, 32, 33, 34], public gathering, and travel restrictions [35, 36, 37] for various countries. Due to the difference in analytical methods and assumptions, the parameter estimates describing COVID-19 dynamics vary widely. This variability is also reflected in the estimates of the effectiveness of public health interventions implemented worldwide. Most epidemiological models of disease transmission are simplistic and use time-invariant transmission rates. However, in reality, due to mitigation efforts and the evolving nature of the infection mechanism, such rates become temporally dynamic. Furthermore, most SIER-type models exclude the effects of testing and subsequent quarantining, and occasionally, even hospitalization. Such practices fail to adequately account for the size of the susceptible population and therefore tend to provide unreliable estimates of the number of asymptomatic persons infected by COVID-19 in the population.

We propose a detailed discrete time semiparametric stochastic dynamic model for COVID-19 spread. The model is expressed through a system of difference equations connecting various interpretable compartments in the disease dynamics such as individuals who are susceptible, asymptomatic but infected, quarantined, hospitalized, dead and have recovered from the disease. We introduce interpretable time-varying parameters to reflect various temporally dynamic rates. Our model also includes available information on the number of tests. On the other hand, the proposed model does not make restrictive and often untestable distributional assumptions about compartments or parameters that are commonplace in various probablistic models for the epidemiological dynamics.

We employ nonlinear nonparametric regression techniques through a profiling based estimation procedure to estimate the model parameters and the number of people in different compartments. Using residual bootstrap based techniques, we also provide point-wise confidence intervals (bands) for the time-invariant (time-varying) parameters. The proposed model and estimation procedure relies on linear kernel weighting and fairly low dimensional optimization, thus avoiding Markov Chain Monte Carlo and other computationally expensive methods employed by Bayesian inference schemes for standard epidemiological models. Therefore, the estimates can be obtained almost instantaneously. Another key feature of our method is the ability of identifying and estimating unobservable quantities such as the actual number of asymptomatic but infected people at any given time. The estimated trajectory of the infected but asymptomatic population over time, its doubling rate, the true case fatality rate, and an analogue of the basic reproduction rate are crucial in interpreting the time-dynamics of the pandemic. They have important implications for policy decisions regarding appropriate mitigation strategies.

## 2. A Multi-compartment Model for Disease spread

We consider a closed population without emmigration or immigration and propose a model for the Covid-19 pandemic spread in terms of various observable and partially or totally unobservable compartments.

Suppose at time *t, C*_*t*_, *D*_*t*_, *T*_*t*_, respectively, denote the number of confirmed cases, number of deaths due to the disease and the number of tests performed up to time *t*. These variables are non-decreasing cumulative counts and are generally fully observed. The number of hospitalised persons due to Covid-19 infection at time *t* (denoted *H*_*t*_) is also generally observed (see Section 3.2 for more detail). Furthermore, we observe *Q*_*t*_, the number of asymptomatic individuals who are in quarantine at time *t*. These individuals have been tested positive, but show no significant symptoms requiring hospitalisation.

The most crucial unobserved compartment is *A*_*t*_, i.e. the number of infected but asymptomatic individuals at time *t*. It is well known that the people in this group are primary spreaders of the disease. Furthermore, due to under-reporting, the number of confirmed cases would be a fraction of *A*_*t*_. Since we do not observe how many in the population are currently infected, the number of susceptible individuals at time *t*, (denoted *S*_*t*_) is also unobserved.

The number of recovered individuals (denoted *R*_*t*_) up to time *t* can be partially observed. To understand this, note that the recoveries from quarantine centres and hospitals, (denoted 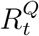 and 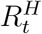 respectively) are reported, though not necessarily separately (see Supplement Section S2., for the case when 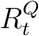 and 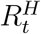 are reported separately). But since *A*_*t*_ is unobserved, the number of asymptomatic but infected people who recover without being quarantined or hospitalised (denoted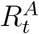) cannot be observed. That is, even though 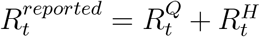 is available from the data, the total recovery *R*_*t*_ is not.

The proposed disease propagation model are based of the following assumptions:

**A1** Only an asymptomatic individual who is not either in quarantine or in hospital can transmit the disease to a susceptible individual.

**A2** People who recover from the disease are immune from subsequent infection.

**A3** False positive rate for the test is negligible, so that if somebody is confirmed to be positive, then he/she is assumed to be infected.

**A4** Anybody who shows significant symptoms, whether being in quarantine or not, is immediately hospitalized, and is tested to be positive.

**A5** There is no effective treatment regime for the asymptomatic individuals, and so they recover or turn symptomatic at the same rate regardless of whether they are tested positive (and hence quarantined) or not.

A graphical representation of the proposed disease propagation model is presented in Figure 1 below. The assumptions A1-A5 are quite general and concur to the observed dynamics of Covid-19 pandemic so far, even though a relatively tiny fraction of people do get infected by prolonged exposure to symptomatic patients, typically in hospitals. However, this small violation of assumption A1 is unlikely to have a significant influence on the overall dynamics, and in any case, the requisite data to account for this violation is practically unavailable. The number of reported reinfection after recovery is negligible, so are the false positive rates of both RTPCR and antigen tests (estimated to be less than 5% [38, 39, 40, 41]). If necessary, the assumptions A2 and A3 can be generalised by adding a fraction of the recovered people in the susceptible category. Assumption A5 implies that the rate of transfer from compartment *A*_*t*_ to 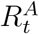 is same as that of transfer from the compartments *Q*_*t*_ to 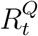 and the rate of transfer from the compartments *A*_*t*_ and *Q*_*t*_ to *H*_*t*_ are equal.

**Figure 1.**
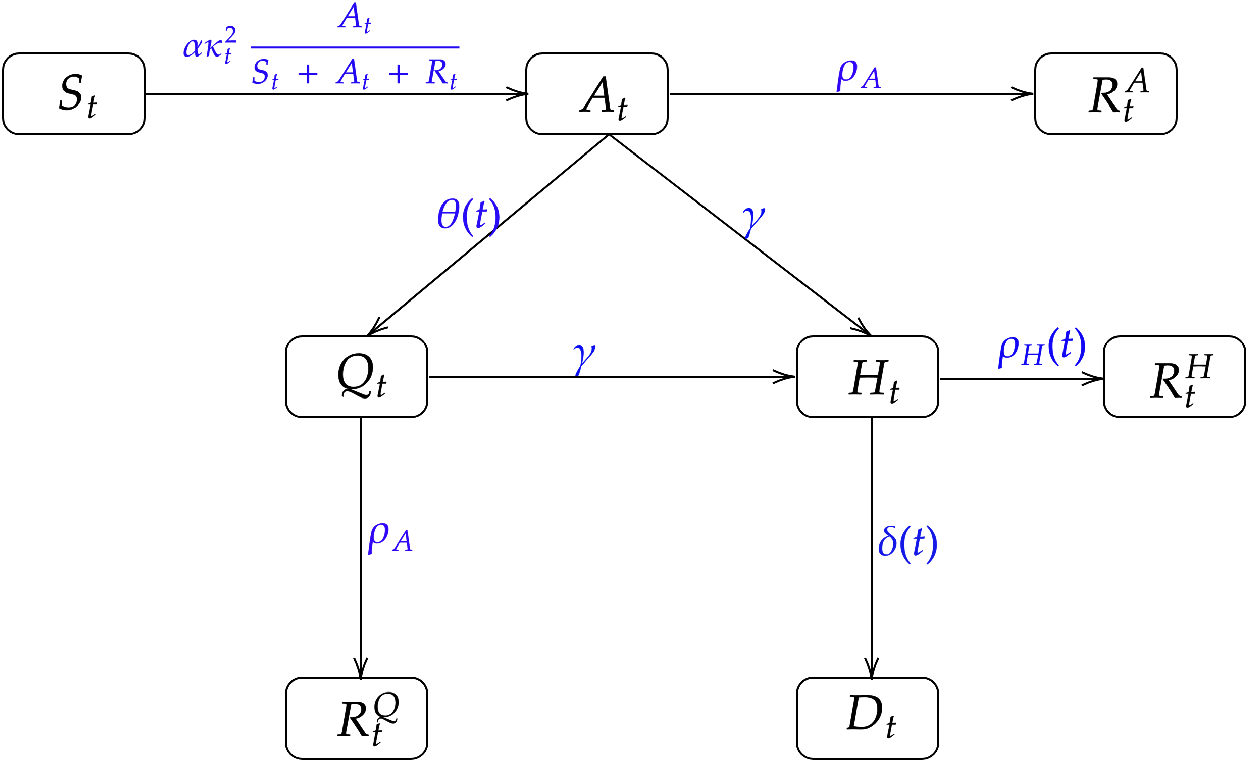
A graphical representation of the disease propagation model. *S*_*t*_, *A*_*t*_, *H*_*t*_, *Q*_*t*_, *D*_*t*_ are the number of susceptible, infected, hospitalized, quarantined, and deceased people at time *t* respectively. 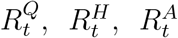 represent the recovered population from quarantined, hospitalization, and infected but asymptotic stages respectively. The rate parameters are as described in Section 2.1.

### 2.1. Disease Propagation Model

We assume an underlying Poisson process model for describing the disease dynamics. Let Δ*C*_*t*_ = *C*_*t*+1_ − *C*_*t*_ be the increments in the number of observed confirmed cases in day *t* + 1. The increments Δ*A*_*t*_, etc. are defined similarly. Under our model, conditionally on the current values of different compartments (collectively denoted by ℱ_*t*_), the above increments follow Poisson distributions with their mean depending on ℱ_*t*_ and a set of rate parameter. Based on our assumptions, the evolution model are expressed as follows:

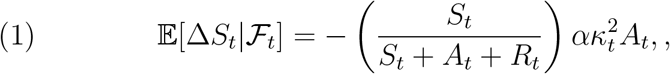

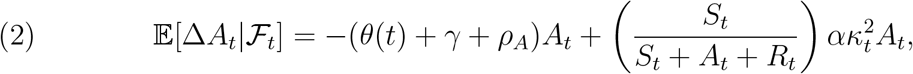

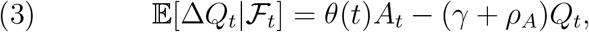

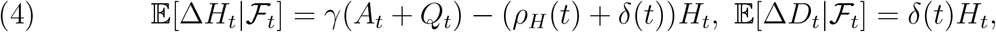

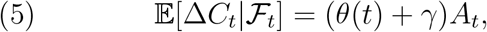

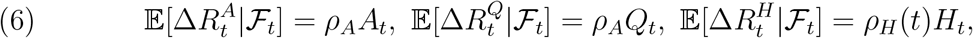

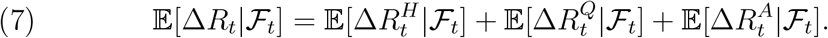

A schematic diagram of the proposed model can be found in Figure 1. All parameters in the proposed model are nonnegative. The parameter *α* is the baseline infection rate, in the absence of any social distancing. This means, *α* is the average number of susceptible individuals who may be infected on any given day by an asymptomatic but infected individual. The rate of daily recovery directly from the asymptomatic compartment is denoted by *ρ*_*A*_. By assumption A5, this is also the daily rate at which a quarantined individual directly recovers. We use *γ* to describe the rate at which an asymptomatic individual may become symptomatic on a given day. By assumption A5, this rate is the same whether the individual is free or in quarantine. The symbols, *ρ*_*H*_(*t*) and *δ*(*t*), respectively, denote is the rate at which people recover and die from the hospitalized compartment. We assume both these rates to be time-varying to reflect the changing levels of effectiveness of treatment regimes over time. We emphasize that Poisson distributions for the increments of various compartments is only a working assumption which guides our estimation strategy (e.g., by formulating appropriate transformations of variables). In Supplement Sections S6. and S7., we carry out a detailed numerical simulation under the Poisson model to validate the statistical performance of the proposed estimation procedure.

Information about daily tests is included in the model using the function *θ*(*t*). We call it the *confirmed fraction* (*CF*), i.e. the fraction of currently asymptomatic individuals who are detected through testing. Parameter *θ*(*t*) would depend on the daily number of tests, as well as the efficiency of the testing strategy in identifying the infected and asymptomatic individuals. The contact tracing strategies were introduced by many states [42, 43] with varying success. In many parts of the world, people in close contact of hospitalised patients are routinely tested. This strategy is closely connected to cluster sampling, where a cluster is defined by the contacts of a hospitalised person. Guided by the above consideration, we reformulate the parameter *θ*(*t*) by expressing it as follows:

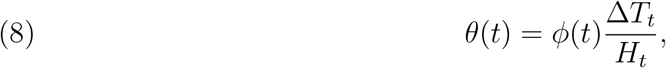

where *ϕ*(*t*) is interpreted as the *testing efficiency* (*TE*) since it measures the fraction of confirmed asymptomatic cases per test, per (currently) hospitalized patient. In Section 5 we estimate *ϕ*(*t*) by minimising certain loss function, from which *θ*(*t*) is subsequently estimated.

In addition, our model (see (1)) depends on a variable *κ*_*t*_, which is the current state of the level of interaction among individuals. Expressed as a fraction, taking value 1 for normal activity, and 0 for complete lockdown, this variable measures the prevalent social distancing in the population. In general *κ*_*t*_ is not observable. However, often a surrogate variable based on data collected by internet service providers such as Google from the usage of smartphones [44] can be used [45, 46, 47].

From (1) the variable 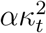 approximately measures the daily rate at which a susceptible individual turns asymptomatic-infected. In the early stage of the epidemic, the fraction *S*_*t*_*/*(*S*_*t*_ +*A*_*t*_ +*R*_*t*_) ≈ 1. Furthermore, rather than waiting for herd immunity to be achieved, mitigation measures are implemented in most affected places or countries to contain the spread of the disease. As a consequence, at any given time, the number of non-susceptible people is much lower as compared to the susceptible population. So *S*_*t*_*/*(*S*_*t*_ + *A*_*t*_ + *R*_*t*_) has remained quite close to 1 for almost the duration of the pandemic until this point, due to the absence of mass-scale vaccination.

Notice that (5), provides a connection between the daily reported confirmed cases Δ*C*_*t*_ and the number of asymptomatic-infected individuals *A*_*t*_ in the population. In our model, an asymptomatic-infected person can be discovered either through a positive test and subsequent quarantining, or through hospitalisation upon showing severe symptoms. Therefore, once the estimates of *θ*(*t*) and *γ* are available, equation(5) allows us to estimate the unknown *A*_*t*_ from the observed *C*_*t*_. It is also clear that, due to unavoidable severe under-reporting, Δ*C*_*t*_ will only be a fraction of the number of total infected individuals at any time point.

### 2.2. Some relevant epidemiological markers

The proposed model is more realistic than the traditional SIR, SEIR etc. models and allows us to estimate different epidemiological markers which can measure the dynamics of disease spread. Our focus here is on estimating epidemiological markers related to the number of asymptomatic but infected persons (i.e. *A*_*t*_) in the population. It is well-known that the disease is mostly spread through persons in that group. Thus the proposed epidemiological markers reveal more fundamental trends of disease dynamics, than what can be obtained only by the confirmed case counts. In particular, we define the following epidemiological markers:

#### 2.2.1. Relative Change in Confirmed Fraction (RCCF)

The relative change in confirmed fraction measures the change in the fraction of currently asymptomatic-infected individuals who are caught in the quarantine net through testing relative to the total fraction of currently infected individuals are either quarantined or hospitalised. From Section 2.1 we get:

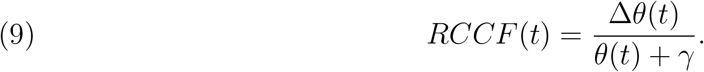

The marker *RCCF* (*t*) measures the dynamics of the efficacy of the testing regime to isolate the asymptomatic but infected individuals from the population into quarantine. From (8), this marker is directly controlled by the prevalent testing strategy and efficiency.

#### 2.2.2. Crude infection rate (CIR) and Net Infection Rate (NIR)

The crude infection rate is defined as the fraction of change in the daily confirmed cases on a day to the number of confirmed cases on that day. In our notation, it follows that:

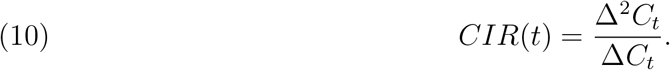

Since CIR suffers from the under-representation inherent in the reported number of confirmed cases, we define a model-based estimate for the infection rate, denoted Net Infection Rate (NIR), which is the ratio of the daily change in the number of asymptomatic-infected persons to the number of the asymptomatic-infected persons. In our notation:

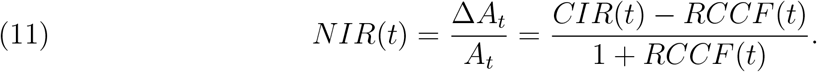

#### 2.2.3. Daily New Infections (NI)

From our model and assumptions, the daily number of new infections are given by the number of susceptible population who turn asymptomatic-infected on that day. From (1) we define this marker as:

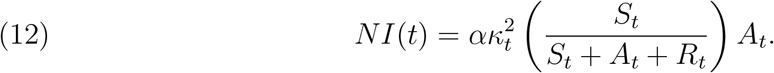

The cumulative number of new infections up to time *t* can be defined as 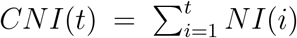

#### 2.2.4. Doubling Times and Rates

The doubling time at time *t*, denoted *t*_*d*_(*t*) measures how much longer it would take for the number of infected upto time *t* to double. The doubling rate at time 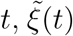 is given by the inverse of the doubling time. A higher doubling rate reflects the faster spread of infection. This rate is often used to measure the effect of social distancing campaigns, improved hygiene and case tracking.

The doubling time for *C*_*t*_ computed using the relationship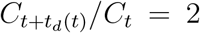. It can be shown (see the Supplement Section S4.) 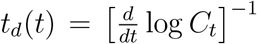. That is the doubling rate 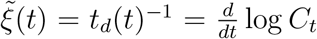. Doubling rates for other compartments can be computed similarly.

#### 2.2.5. Crude and Net Case Fatality Rates

In general a *case fatality rate* at time *t* is given by the ratio of the total death count and the total case count at that time. Depending on whether the reported case counts or the actual case counts are used, we can define two different case fatality rates. The *crude case fatality rate* (CFR) is defined as

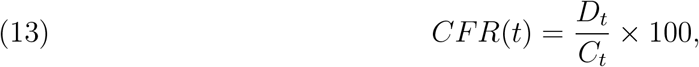

whereas the *net case fatality rate* is given by

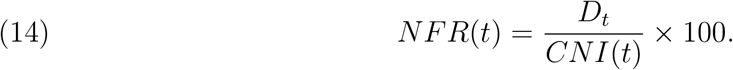

#### 2.2.6. The Basic Reproduction Rate

In the conventional SIR or SIER models, basic reproduction rate (*R*_0_), which measures the expected number of cases directly generated by one case in a population where all individuals are susceptible to infection [48] is used to determine the nature and rate of growth of the pandemic. Unlike these models, our model is more detailed and allows for time varying parameters and as a result, the conventional *R*_0_ cannot be directly estimated from our model. The closest epidemiological quantity we can observe is the background infection rate, *α*, measuring the average number of susceptible individuals who may be infected on any given day by an asymptomatic but infected individual. However, by following the next generation method [49, 50] an analogue of the basic reproduction rate for the compartment *A*_*t*_ can be computed.

By focusing on the compartment *A*_*t*_, under our assumptions from (2) new infections arrive at the compartment at the rate of 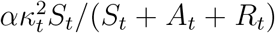 and leave at the rate of (*θ*(*t*) + *γ* + *ρ*_*A*_). There is no other pathway for disease spread. Thus we can define an analogue of the basic reproduction rate as:

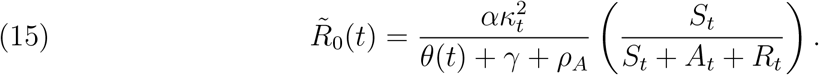

Note that, the proposed 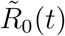 can be interpreted in the same way as the conventional basic reproduction rate. By construction 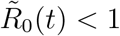 indicates negative growth of the number of asymptomatic-infected persons, whereas 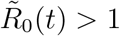 indicates its positive growth. However, temporal variation of 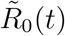 is more complex. Assuming that,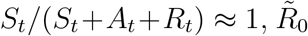 can decrease with time either due to reduction in *κ*_*t*_, that is the current state of interaction among individuals, or due to an increase in the confirmed fraction *θ*(*t*). That is, the proposed 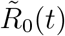 is directly influenced by the mitigation efforts such as social distancing, adherence to use of masks, increased testing and subsequent quarantining, hospitalisation of symptomatic patients etc.

Most epidemiological models such as SIR, SIER etc., assume fixed doubling rate parameters. In reality, however, the doubling time is a dynamic quantity, which changes continuously due to mitigation efforts and the inherently changing nature of virus-spreading mechanisms. It is then vital that policymakers and researchers have access to frequent and up-to-date estimates of doubling time [51]. For example, [52] provided early, fixed-in-time estimates of epidemic parameters of COVID-19 (e.g. growth rate, doubling time, basic reproduction number, case detection rate) during the first 50 days of onset in China. In recent work [53, 54] the basic reproduction number and doubling time have been studied in a dynamic manner by considering a varying coefficient model with daily new cases as the response and time as a predictor. A related approach focused on the real-time estimation of case fatality rates using Poisson mixture models can be found in [55].

## 3. Results: Application to COVID-19 data from the USA

### 3.1. Data Preparation

We consider the dynamics of the spread of COVID-19 in various states of USA for a tentative time window of late April to mid December. The proposed model is based on the observed state-wise daily counts of confirmed infections, deaths, hospitalisations and reported recoveries from the hospitals and quarantining facilities. Daily counts of the confirmed COVID-19 cases in various states were obtained from the COVID-19 Data Repository maintained by the Center for Systems Science and Engineering (CSSE) at Johns Hopkins University. This is publicly available at https://github.com/CSSEGISandData/COVID-19 and was accessed on December 15, 2020. The state-wise daily counts of positive and negative COVID-19 test results, current hospitalization, and recovery per day and state, were obtained from the CDC data repository - the COVID Tracking Project and are publicly available at https://COVIDtracking.com/ (accessed on December 15, 2020.)

The collected noisy data used is pre-processed and cleaned, removing the irregularities present in the recording and maintenance of the data repositories. Any missing or evidently wrong (e.g. negative counts) observations were replaced by the average of the data from adjacent five days. Inherent noise present in the daily counts was removed by pre-smoothing the trajectories using a *Lowess* method [56, 57, 58, 59] with bandwidth 1*/*16.

### 3.2. Results

Unfortunately, a continuous record on hospitalization and recovery information were not available for many states. For example, most counties in California are not reporting recovery information. Data on Hospitalization is found to be updated once a week in Massachusetts and Florida. New York on the other hand started documenting the hospitalization information only after the initial surge of the pandemic was over for the state. In our analysis we only consider the states for which daily observations on 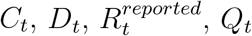, and *H*_*t*_ are available throughout the time window under consideration. Any missing/negative values are replaced by the average of the adjacent five days’ data. For a few states e.g. Alabama, the available data turned out to be too unreliable. We present results for fifteen states in US which demonstrate the efficacy of the proposed model and the estimation methods. For succinct representation, the results from only one state i.e. Utah is presented in details below. The results for other fourteen states can be found in the Supplement Section S9.

#### 3.2.1. Case study for the state Utah

We present our results for the state *Utah* for the time window between 7th May, 2020 to 4th December, 2020. The time interval includes the Thanksgiving weekend (27th −28th November, 2020), when due to the long holiday, the reported data may be unreliable. In Figure 2 plots of various time-varying compartments and epidemiological markers defined in Section 2.2. The plots of the parameters with their residual bootstrap confidence intervals can be found in Figure 4. Due to unreliable reporting around the Thanksgiving holiday, the estimates values after 21st November, 2020 should be interpreted with caution.

**Figure 2.**
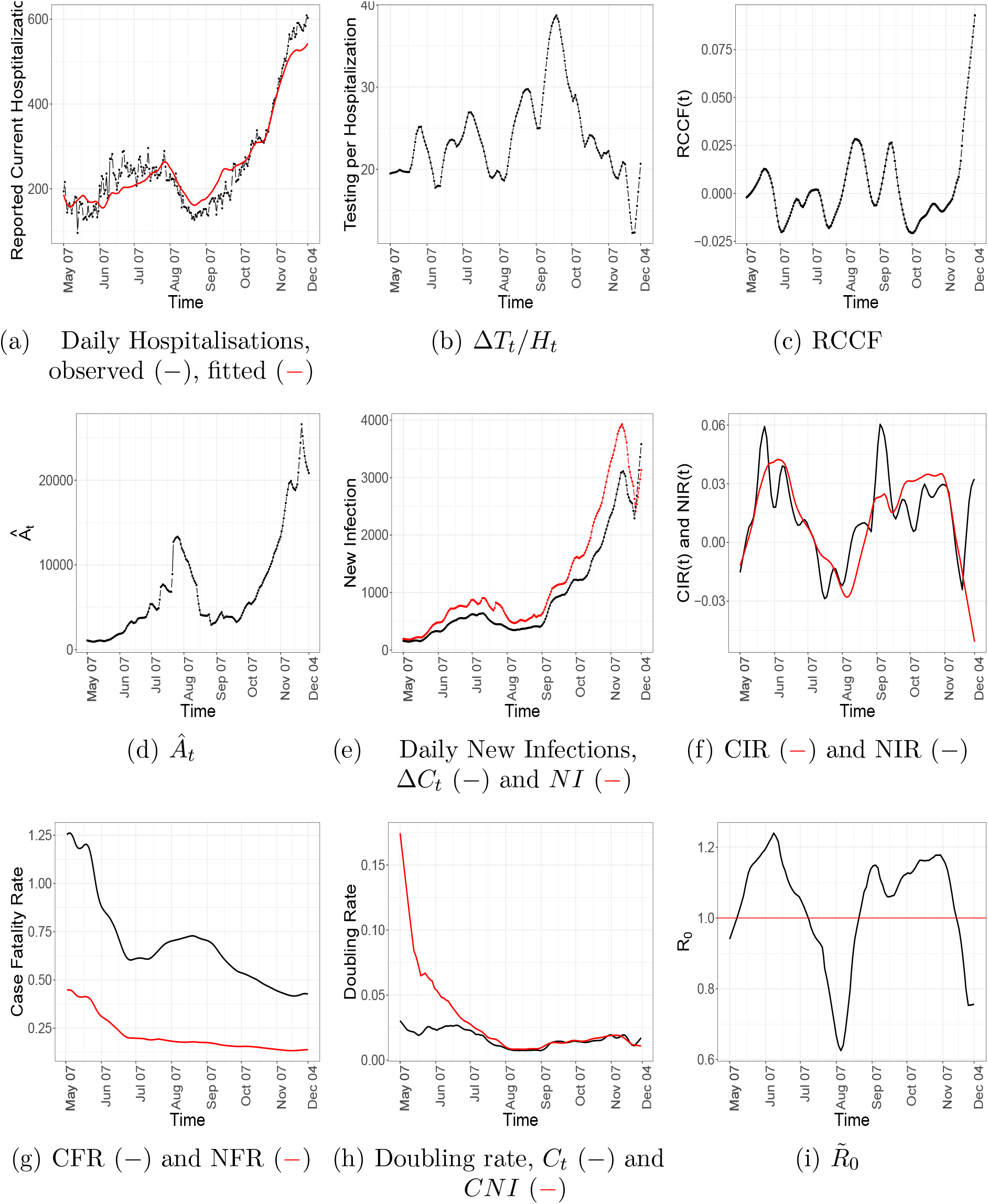
Temporal patterns of some compartments and epidemiological markers for Utah.

**Figure 3.**
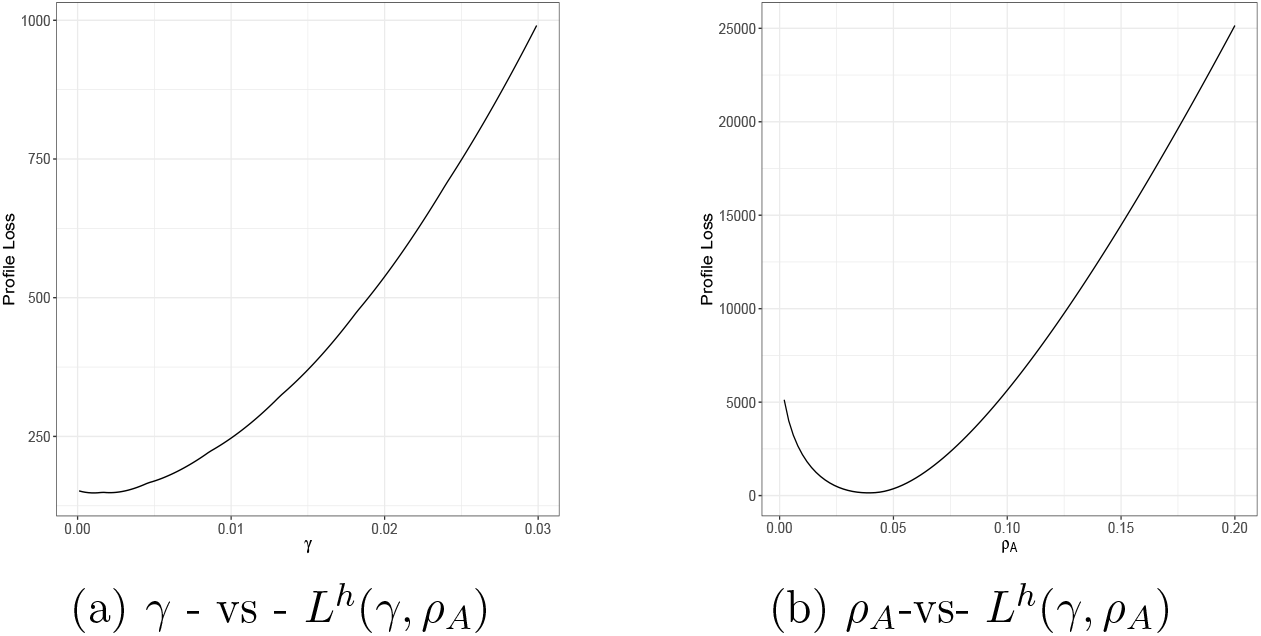
The profile loss as a function of *γ* and *ρ*_*A*_ respectively.

**Figure 4.**
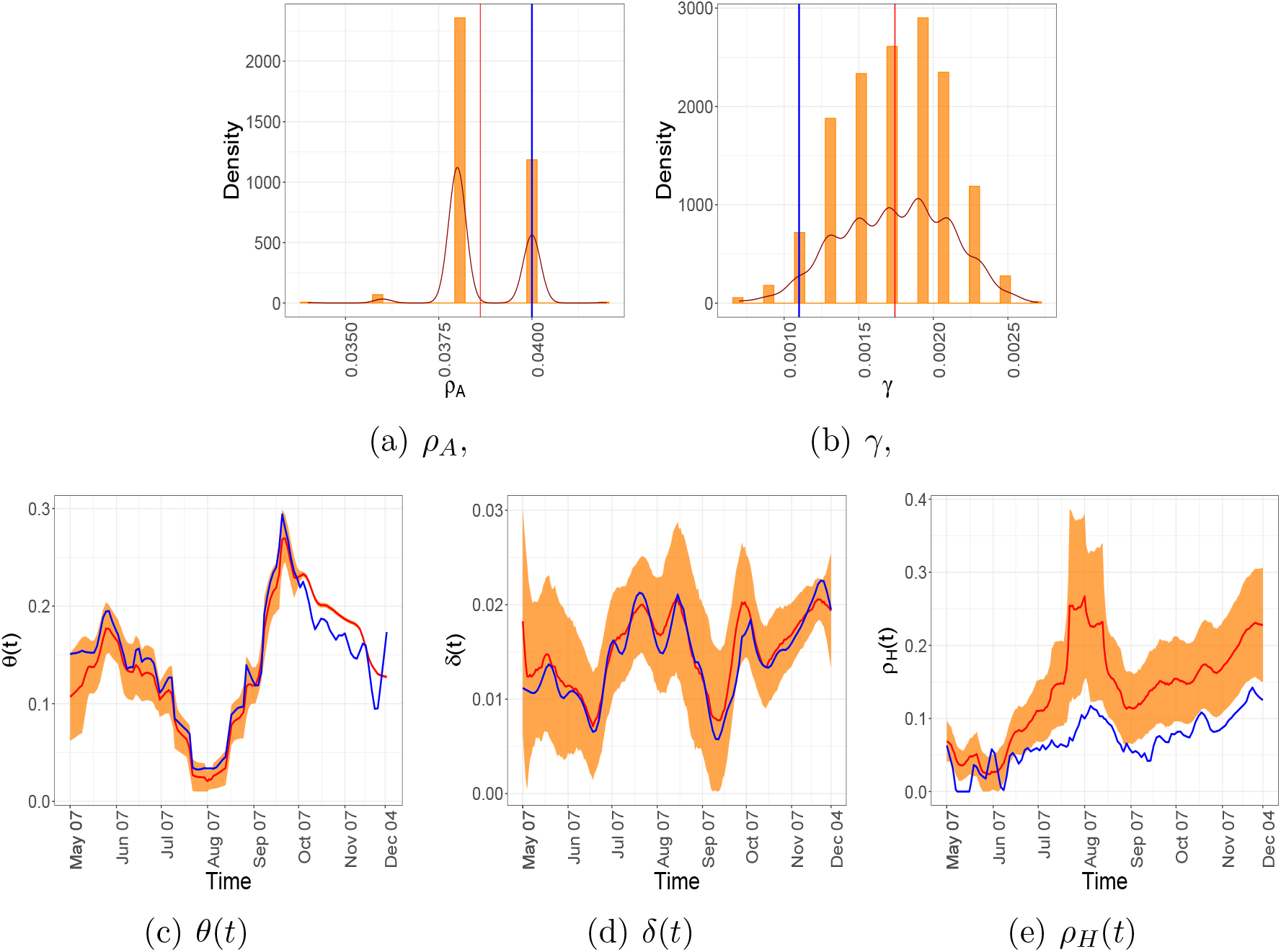
Estimates and residual bootstrap based confidence intervals for time invariant and time-varying parameters for the state of Utah. The estimate from the data is in blue. The 95% confidence band is in yellow and the mean of the bootstrap estimates are presented in red.

The curves in Figure 2(a) compare the observed and the fitted number of daily number of people in the hospitals. It can be seen that, the fitted values obtained from the model closely follow the observed values. This validates our proposed model and the estimation procedure. From the data and the fit two waves of infection can be identified. It seems the first wave starts at the end of May, 2020 stabilises and begins to die down around 7th August, 2020. The daily number of people in hospitals starts increasing again around the end of August, 2020.

The estimated number of infected asymptomatic people (Figure 2(d)) shows a similar pattern. From a high point around the beginning of August it dips to a low value at the end of August. The number remains stable for a few weeks and starts growing again at the end of September. The two waves can also be clearly observed from the plot of the proposed analogue of the basic reproduction rate 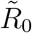 in Figure 2(i). In the time period under consideration, the estimated 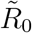 was larger than one in two sub-intervals, namely from middle May to middle of July and then from end of August to beginning of November. The plot of the number of daily new and daily reported infections (Figure 2(e)) shows a local maxima near the middle of November. However, we cannot rule out the boundary effect as its cause.

The plots of CIR and NIR seem to be similar (Figure 2(f)). In fact, the observed doubling rate obtained from *C*_*t*_ and that estimated from *CNI* seems to be very close in the second wave of the pandemic (see Figure 2(h)). This implies that in the second wave the reporting kept pace with the spread of the disease. Figure 2(g) shows the crude and net fatality rates. Due to the denominator effect, naturally the crude fatality rate is much faster than the net fatality rate. However, our estimate of NFR is mostly below 0.25%, which complies with widely held beliefs [60, 24, 61, 62].

The estimate of *δ*(*t*) in Figure 4(d) seems to remain stable throughout the time period under consideration. The 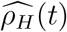 shows an overall increasing trend. On the other hand, The estimate of *θ*(*t*) decreases to a near zero value at the end of the first wave (7th August, 2020) it then increases to its maximum value at the end of September and starts to decrease again. The parameters (*γ, ρ*_*A*_) are estimated based on minimization of the profile loss using a grid search algorithm with grid size 0.0001. In Figures 4(a) and 4(b) the estimates from residual bootstrap samples take discrete values, resulting in a discrete histogram counts.

The daily number of tests and its effect in quarantining asymptomatic but infected people can be judged from the Figures 2(b) and 2(c). The state of Utah increased its testing capacity by public-private partnership. Empirical comparison of the Figures 2(a) and 2(b) seems to reveal that although the number of daily tests could keep pace with daily number of hospitalised patients up to the third week of September, but growing number of hospitalised people ultimately outpaced the number of daily tests. Note that, estimated *θ*(*t*) increases at the onset of the second wave (see Figure 4(c) between 7th, August and 21st, September), however, from figure 2(d), *Â*_*t*_ remains more or less constant. Thus, growth in the number of new infections could be due to the increase in *κ*_*t*_, that is due to more interaction among individuals and less social distancing.

#### 3.2.2. Summary of Results for Other States

We present a summary of the results obtained from applications of proposed method on the data procured from fifteen other states in USA. The estimated parameters are in Table 2. The time-varying parameters, (*ϕ*(*t*), *ρ*_*H*_(*t*), *δ*(*t*)), are summarised by their means. The computed 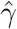, that is, the rate for an asymptomatic person turning symptomatic on a particular day is the smallest in Arizona and largest in Tennessee. This estimate is smaller than 0.001 for Arizona and Idaho. Minnesota, has by far the highest recovery rate for an asymptomatic person without needing hospitalization on a particular day (i.e. 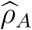). For Iowa, Nebraska, Pennsylvania, and Utah this rate is comparable and reasonably high, whereas Arizona, Delaware and Idaho have their 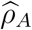 value below 0.01. The average confirmed fraction 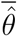 is larger than 0.1 in Delaware, Tennessee and Utah. It is lowest in Texas. This can be associated with better estimates obtained for these states due to the availability of more reliable data, whereas for Idaho, South Dakota, and Texas, a lower value of there epi-markers tend to give evidence for a more relaxed testing paradigm. More testing is required for isolating the confirmed cases to contain the disease faster, which can be reflected in the numbers for these states. The detailed results and bootstrap confidence regions for these additional states can be found in the Supplement Section S9.

**Table 1.** Estimates, and the residual bootstrap Confidence intervals, mean and standard deviations for the time-invariant parameters for Utah. The latter three are computed based on 1000 bootstrap resamples.

|  | Estimate | 95% Confidence Interval | Mean | s.d. |
| --- | --- | --- | --- | --- |
| $\gamma$ | 0.0011 | [0.0011, 0.0023] | 0.0017 | 0.0003 |
| $\rho_A$ | 0.0400 | [0.0380, 0.0400] | 0.0386 | 0.0010 |

**Table 2.**
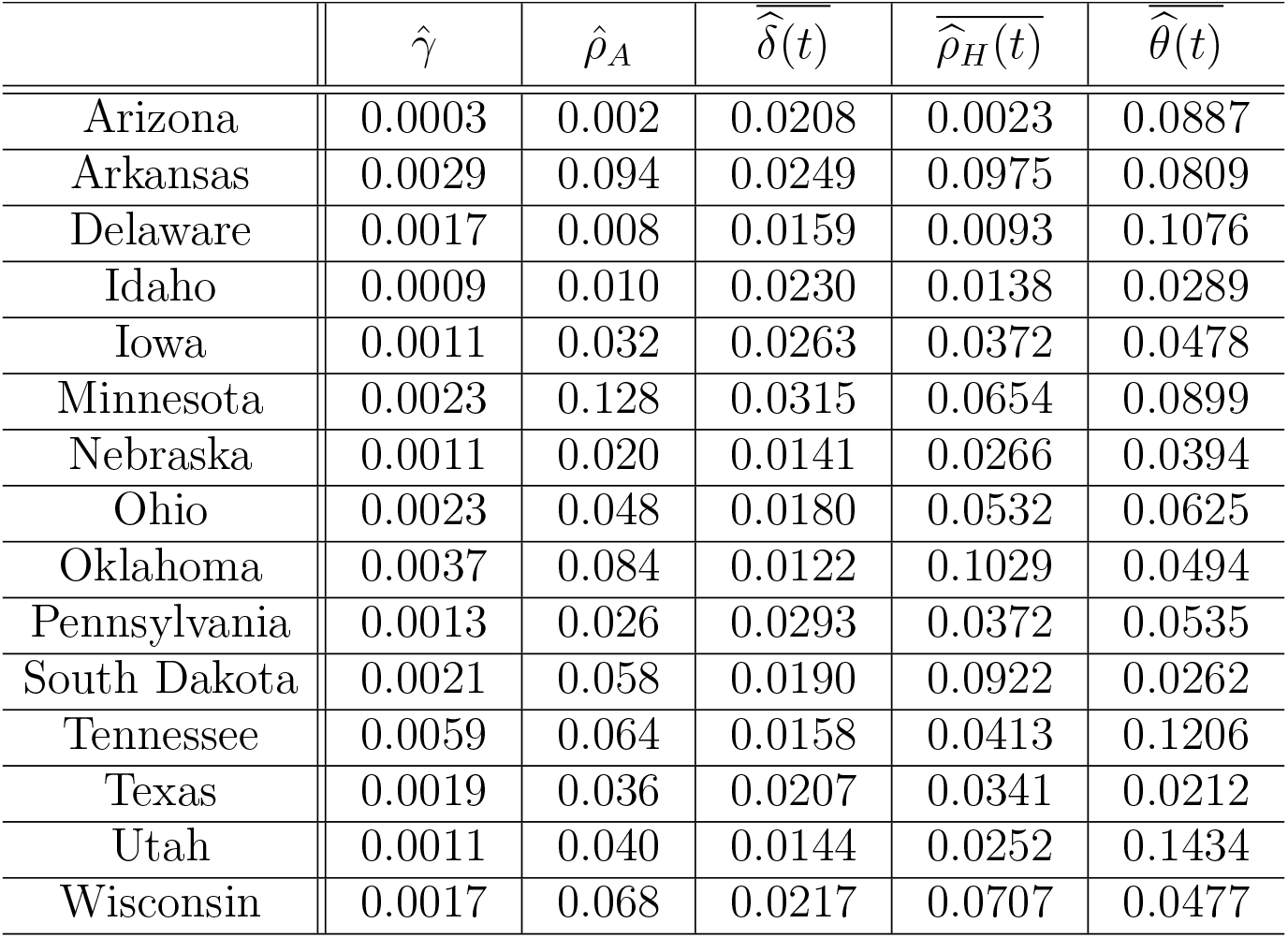
Mean estimated parameters for different states in the USA.

Among the states not included in Table 2, many, such as California did not report all the required compartments. For many states such Alabama, Colorado, Maryland, Massachusetts, North Carolina etc. the reported data produced monotone profile likelihoods which yielded unreliable boundary estimates. This could be due to the change in definition of many compartments over time, which violated our assumptions. Furthermore, for some states such as New York, New Jersey, Michigan etc., the pandemic started quite early and ran its course even before a proper testing protocol and other mitigation measures could be introduced. Thus the data from these states is contaminated with an inherent bias, the number of people in quarantine or symptomatic states are to too low to produce reliable estimates.

## 4. Discussion

We introduce a multi-compartment model for COVID-19 dynamics which can incorporate data from compartments like quarantine, hospitalisation etc. Unlike the conventional SIR and similar models, the proposed model is based on interpretable time-varying parameters, which are more suitable for describing the disease dynamics in the presence of mitigating procedures. It also incorporates the information about testing and subsequent quarantining. We estimate the model parameters using profile likelihood and nonparametric regression. This provides a much faster alternative to Markov Chain Monte Carlo based Bayesian models which are commonly used in estimating SIR parameters. Using the proposed detailed and robust model one can estimate the daily number of asymptomatic but infected individuals, who are universally regarded as the key agent for the COVID-19 spread. We define several epidemiological markers which uses the number of asymptomatic-infected individuals and therefore reveal the true underlying dynamics of the pandemic.

Our model only uses information on the number of confirmed infected, hospitalised, deaths and total reported recoveries from hospitals and the quarantine. We don’t require those numbers separately. However, such numbers are often available. In such a case, the loss function in (21) can be simplified a bit. The details can be found in the Supplement in Section S2.

The model parameters have been estimated assuming that no information about the mobility within the population is available. Such information identifies the parameters *κ*_*t*_ and *α* in our model. Reliable data on the compliance to social distancing, mask wearing etc. are difficult to get. Various aspects of the mobility data available from Google can be one potential surrogate for *κ*_*t*_ [63, 64]. However, such data only look into the fraction of people going to workplace or recreation and so on, and does not collect information on the people who are the super spreaders or not wearing masks. Thus, it does not necessarily reflect the the social mobility index *κ*_*t*_, as incorporated in our model. In the Supplement (see Sections S3. and S9.), we present results by using the Google mobility data as a surrogate to *κ*_*t*_. In particular, information on the change in the mobility patterns, as the percentage decrease (increase) from the baseline, in different areas such as parks, residential locations, retail locations, among others during the pandemic from the Google mobility database were obtained. The publicly available data was sourced from https://www.google.com/COVID19/mobility/ (accessed on December 15, 2020). When information on *κ*_*t*_ is available, the parameter *α*, which is the average number of susceptible individuals who may be infected in a day by an asymptomatic-infected individual is identifiable and can be estimated. The details can be found in the Supplement Section S3.

The proposed method and estimation procedure do not explicitly use the underlying assumption of a Poisson process. In the Supplement (see Section S6.∓S8.), however, we use an ensemble of independent Poisson processes to simulate data from the proposed model. These aggregated data sets are then used to accurately estimate various parameters, which validate our estimation procedure. The aggregation has the effect of increasing the number of observations in the compartments and thereby improving estimation accuracy. If the number of individuals in the symptomatic or quarantined compartments are low, e.g. at the onset of the pandemic, inherent biases are introduced in the estimated trajectories. A bigger sample size is required to correct such contaminants.

It should be noted that COVID-19 analyses based on published case and death counts, including those conducted here, are subject to the same biases which affect the accuracy of the data, primarily due to under-reporting [65] or mis-recording of the data, the degree of which varies by country [66]. The reasons for such under-reporting are many, including insufficient testing materials, political incentives, and administrative delays. Furthermore, our model assumes a closed population. It ignores migration between cities, states or countries which play an essential role in the propagation of the disease. We only count the deaths solely due to Covid-19 infections and as such completely ignore any competing causes of morbidity, as well as increase in population due to new births.

With this caveat in mind, the study of available data presented in this article nevertheless provides useful insights into the COVID-19 propagation and ways to control it. It clearly follows that in order to break the chain of transmission and “flatten the curve”, we need extensive testing and adhere to strict social distancing protocols.

## 5. Methods : Parameter and compartment Estimation

The core of our estimation strategy is to utilize (1)–(7) to formulate appropriate regression problems. Our estimation procedure is based on the availability of the compartments *C*_*t*_, *D*_*t*_, *H*_*t*_, *Q*_*t*_, *T*_*t*_ and 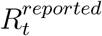 only. We do not assume that data on the social distancing factor *κ*_*t*_ is available. Described crudely, the proposed estimation method uses local regression (linear or nonlinear) methods for estimating the time-varying parameters, while *profiling* over the time-independent ones.

In the absence of data on *κ*_*t*_, the parameter *α* in (1) is not identifiable. We first describe how the product 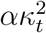 can be estimated. Notice that, ignoring the stochasticity, we may rewrite (5) as

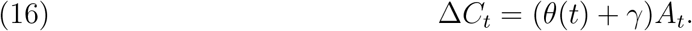

Defining *η*(*t*) = *θ*(*t*) + *γ*, and applying the difference operator on both sides of (16), and finally dividing both sides by Δ*C*_*t*_, we obtain

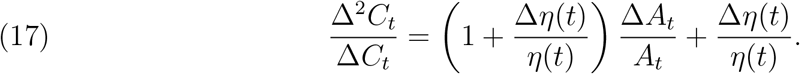

Now, ignoring the second order factor (Δ*η*(*t*)Δ*A*_*t*_)*/*(*η*(*t*)*A*_*t*_), from (2), at the onset of the epidemic (i.e. *S*_*t*_*/*(*S*_*t*_ + *A*_*t*_ + *R*_*t*_) ≈ 1), we have the approximate relationship:

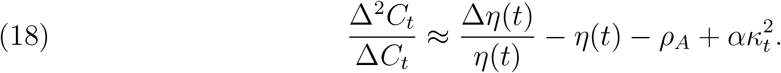

Note that (18) establishes an approximate linear relationship, between the observable quantity Δ^2^*C*_*t*_/*C*_*t*_ and the product 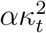. Below we show that, the other parameters in (18) can be estimated, from the available data. These estimates can be plugged in to get an estimate of 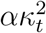.

### 5.1 Point Estimates

Broadly speaking, the estimation strategy consists of separating the time-dependent and time-independent parameters, into vectors ***β***_*t*_ = (*ϕ*(*t*), *ρ*_*H*_(*t*), *δ*(*t*)) and ***ζ*** = (*γ, ρ*_*A*_) respectively. First the vector ***ζ*** is kept fixed and for each *t* the time dependent parameter ***β***_*t*_ is estimated (denoted 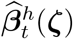) by minimizing the “conditional” local loss function 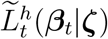 (described below) with respect to ***β***_*t*_, subject to appropriate constraints on the parameters (non-negativity as well as certain upper bounds). The optimal local conditional loss is then combined across different time points to obtain the *profile loss* function for ***ζ***, which is given by

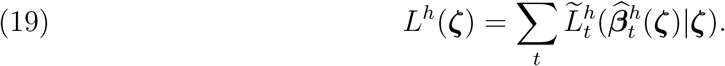

The estimate 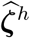 of ***ζ*** is obtained by minimizing *L*^*h*^(***ζ***) under appropriate constraints. We update the estimates of ***β***_*t*_ as 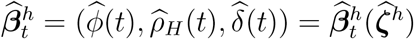.

In order to define the conditional loss function, let *K*(·) be a nonnegative kernel integrating to one. Now, for a bandwidth parameter *h* > 0, the *local weighted conditional loss* function of ***β***_*t*_, given ***ζ*** is defined as:

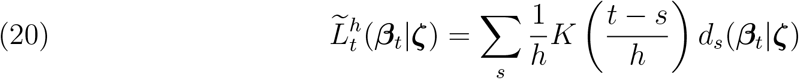

Where

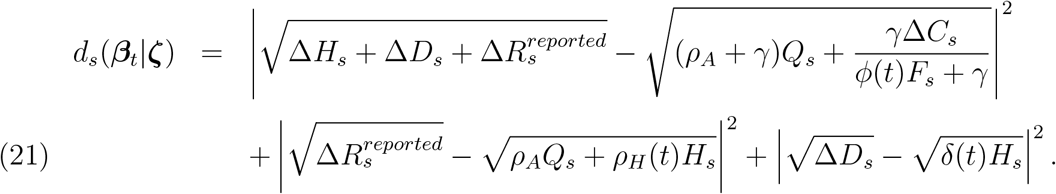

Note that the RHS of (21) only uses the observed data. The first addendum originates from equations (4), (5) and (6). The second and the third term use equations (6) and (4) respectively. The square-root transformation of the responses are used as a variance stabilising transformation, which is driven by the assumed Poissonian characteristics of the responses. Also by construction, the estimate of *δ*(*t*) does not depend on ***ζ***.

Estimated values of the parameters readily yields estimates of the key compartments of the model. In particular, from the definition of *θ*(*t*), (16) and (17) we get:

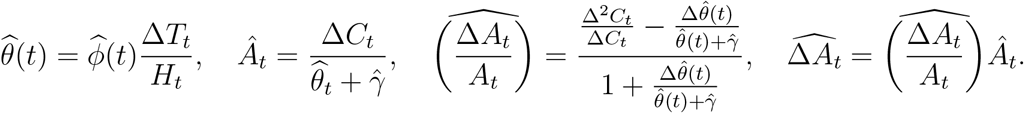

Now, by plugging in 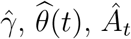 and 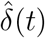 in (4) we get an updated estimator of *ρ*_*H*_(*t*) as

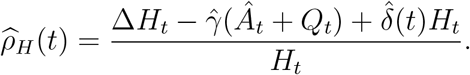

Finally, using (17) an estimate of 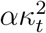 can be obtained as:

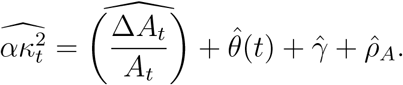

The rest of the compartments can be estimated by plugging in the appropriate parameter or compartment estimates in equations (1) – (7) (see the Supplement Sections S1. and S3.).

The tuning parameter *h* in (20) is obtained by minimising a standardised *L*_1_ distance between the fitted and model based estimates of various compartments through a cross-validation strategy. The actual minimisation is achieved by a grid-search. Details can be found in the Supplement Sections S1. and S2.

### 5.2. Confidence Intervals

We employ residual bootstrap [67, 68, 69] to compute the confidence intervals for our parameter and compartment estimates. Briefly put, the technique adds resampled residuals to the fitted values to create several “resampled” datasets. The point estimation technique described above is applied to each of these resampled datasets to create a new set of parameter and compartment estimates. The empirical distribution of these estimates are then used to construct the confidence interval. The details of the algorithm can be found in the Supplement Section S5. The theoretical validity of the residual bootstrap method is well justified in existing literature [70, 71].

## Data Availability

All data necessary for the replication of our results are collated in https://github.com/Satarupa3671/COVID-19-Nonparametric-Inference.

The data for the number of COVID cases, deaths, hospitalizations, and recovery were originally collected from https://covidtracking.com/data/download while the social mobility data was sourced from https://www.google.com/covid19/mobility.

https://github.com/Satarupa3671/COVID-19-Nonparametric-Inference

## 6. Data and Code Availability

All data necessary for the replication of our results is collated in https://github.com/Satarupa3671/COVID-19-Nonparametric-Inference. The data for the number of COVID cases, deaths, hospitalizations and recovery were originally collected from https://covidtracking.com/data/download while the social mobility data was sourced from https://www.google.com/covid19/mobility.

All code necessary for the replication of our results is collated in https://github.com/Satarupa3671/COVID-19-Nonparametric-Inference.

