## Supplemental information for "Inference on the dynamics of the COVID pandemic from observational data"

#### S1. Choice of bandwidth $h$

The optimization of the profile loss function and hence the parameter estimation depends on the choice of the tuning parameter $h$. We choose the optimal $h$, that minimizes the standardized $L_1$ distance between the fitted and model based estimates for (i) the new number confirmed cases at time $t$, (ii) the number of new infections at time $t$, and (iii) the number of cumulative infections at time $t$.

It should be noted that the number of new confirmed cases at time $t$ (denoted $C_t$) is an observable quantity and often under-reported due to the prevalence of the asymptomatic population. The comparable model-based new infection is approximately given by $\widehat{\alpha\kappa_t^2}\widehat{A}_t$. This model-based estimate depends on the choice of $h$. The difference between the above two quantities measures the number of unreported new cases.

The number of new asymptomatic infections can be calculated in two ways- the model-based growth in $A_t$ given by $\widehat{\Delta A_t} = NIR(t)A_t$ (see Section 2.2.2 for the definition of

$NIR(t)$), and the difference of the estimated $A_t$ fitted from the data given by $\Delta\widehat{A_t}$. A higher value of the discrepancy between the two quantities is likely to indicate a model mismatch. To this end, the cumulative version of the above difference is also considered.

The three different objective functions contribute to the efficacy of the model in the context of the observed data. Combining these in an additive manner, we then proceed to find the minimizer $h$. As such,

$$h^* = \underset{h}{\operatorname{argmin}} \sum_t \left( \frac{|\widehat{\alpha\kappa_t^2}(h)\widehat{A_t}(h) - \Delta C_t(h)|}{|\widehat{\alpha\kappa_t^2}(h)\widehat{A_t}(h)| + |\Delta C_t(h)|} + \frac{|\widehat{\Delta A_t}(h) - \Delta\widehat{A_t}(h)|}{|\widehat{\Delta A_t}(h)| + |\Delta\widehat{A_t}(h)|} \right.$$

$$\left. + \frac{|\sum_{s=1}^t \widehat{\alpha\kappa_s^2}(h)\widehat{A_s}(h) - C_t(h)|}{|\sum_{s=1}^t \widehat{\alpha\kappa_s^2}(h)\widehat{A_s}(h)| + |\Delta C_t(h)|} \right) \tag{S.1}$$

#### S2. A Different Scenario of Reported recovery

The estimation procedure may differ in case $R_t^H$ and $R_t^Q$ were reported separately. In such case, we give a different version of the loss $d_s(\boldsymbol{\beta}(t)|\boldsymbol{\zeta})$ below, corresponding to this alternate reporting regime.

$$d_s(\boldsymbol{\beta}(t)|\boldsymbol{\zeta}) = \left| \left(\Delta H_s + \Delta D_s + \Delta R_s^H\right)^{1/2} - \left(\gamma Q_s + \frac{\gamma \Delta C_s}{\phi(t)F_s + \gamma}\right)^{1/2} \right|^2$$

$$+ \left| \sqrt{\Delta R_s^Q} - \sqrt{\rho_H(t)}\sqrt{H_s} \right|^2 + \left| \sqrt{\Delta D_s} - \sqrt{\delta(t)}\sqrt{H_s} \right|^2. \tag{S.2}$$

In either (21) (see Section 5.1) or (S.2), the use of square-root transformation of the response is driven by the Poissonian character of the responses, and is done to stabilize the variance of the noise.

The estimation strategy consists of first fixing $\boldsymbol{\zeta} = (\gamma, \rho_A)$ and estimating $\boldsymbol{\beta}(t) = (\phi(t), \rho_H(t), \delta(t))$ for each $t$ by minimizing the "conditional" loss function $\widetilde{L}_t^h(\boldsymbol{\beta}(t)|\boldsymbol{\zeta})$ with respect to $\boldsymbol{\beta}(t)$, subject to appropriate constraints on the parameters (non-negativity as well as certain upper bounds). Let us denote the resulting estimate by $\widehat{\boldsymbol{\beta}}_h(t|\boldsymbol{\zeta})$. Notice however that the estimate of $\delta(t)$ does not depend on $\boldsymbol{\zeta}$, while if we use the description (S.2) for $d_s(\boldsymbol{\beta}(t)|\boldsymbol{\zeta})$, then the estimate of $\rho_H(t)$ also does not depend on $\boldsymbol{\zeta}$. After this, we combine the minimum value of the local loss across different time points to obtain the *profile loss* function for $\boldsymbol{\zeta}$ given by

$$L^h(\boldsymbol{\zeta}) = \sum_t \widetilde{L}_t^h(\widehat{\boldsymbol{\beta}}_h(t|\boldsymbol{\zeta})|\boldsymbol{\zeta}). \tag{S.3}$$

We obtain estimates $\widehat{\boldsymbol{\zeta}}_h$ by minimizing $L^h(\boldsymbol{\zeta})$ under appropriate constraints on $\boldsymbol{\zeta}$. Finally, the final estimates of $\boldsymbol{\beta}(t) = (\phi(t), \rho_H(t), \delta(t))$ are obtained as

$$\widehat{\boldsymbol{\beta}}_h(t) = (\widehat{\phi(t)}, \widehat{\rho_H(t)}, \widehat{\delta(t)}) = \widehat{\boldsymbol{\beta}}_h(t|\widehat{\boldsymbol{\zeta}}_h).$$

#### S3. Estimation when $\kappa_t$ is observed

We can adapt an iterative estimation method incorporating the information available on the social mobility parameter $\kappa_t$. The algorithm can be summarized as follows.

- Plug in the values of $\widehat{\eta}(t)$ in (18) (see Section 5) to create a new response $(\Delta^2 C_t/\Delta C_t) - \Delta\widehat{\eta}(t)/\widehat{\eta}(t) + \widehat{\eta}(t)$ and regress linearly onto $\kappa_t^2$.
- The slope gives a (crude) estimate of $\alpha$. Negative of the intercept gives a new estimate of $\rho_A$. Plug in the new estimate $\widehat{\rho}_A(1)$ in the profile loss and minimize $L^h(\gamma, \widehat{\rho}_A(1))$ to obtain new estimates $(\widehat{\gamma}^{(2)}, \widehat{\boldsymbol{\beta}}^{(2)}(t))$.
- Iterate until convergence of the parameters.

We can refine the estimation strategy further by replacing the predictor $\kappa_t^2$ by $\left(\frac{\widehat{S}_t}{\widehat{S}_t+\widehat{A}_t+\widehat{R}_t}\right)\kappa_t^2$ in the regression step, where $\widehat{S}_t$, $\widehat{A}_t$ and $\widehat{R}_t$ are the (model-based) fitted values of the respective states based on the current estimates of the parameters.

$$\widehat{\theta(t)} = \widehat{\phi(t)} \frac{\Delta T_t}{H_t}; \quad \hat{A}_t = \frac{\Delta C_t}{\widehat{\theta(t)} + \hat{\gamma}} \tag{S.4}$$

$$\widehat{\Delta R_t^H} = \widehat{\rho_H(t)} H_t; \quad \widehat{\Delta R_t^Q} = \hat{\rho}_A \; Q_t; \quad \widehat{\Delta R_t^A} = \hat{\rho}_A \widehat{A_t}, \tag{S.5}$$

$$\widehat{\Delta R_t} = \widehat{\Delta R_t^H} + \widehat{\Delta R_t^Q} + \widehat{\Delta R_t^A}; \quad \widehat{\Delta S_t} = \widehat{\alpha \kappa_t^2} \; \frac{\hat{S}_t}{(\hat{S}_t + \hat{R}_t + \widehat{A_t})}, \tag{S.6}$$

where $\widehat{S}_t$, $\widehat{A}_t$ and $\widehat{R}_t$ are the iteratively fitted values of the respective states based on the current estimates of the parameters.

The baseline infection rate in the absence of any social distancing, given by $\alpha$, can also be explicitly estimated in this case using information on the observable mobility data $\kappa(t)$ as follows

$$\hat{\alpha} = \frac{\sum_{t=1}^{T} \widehat{\alpha \kappa_t}}{\sum_{t=1}^{T} \widehat{\kappa}_t}. \tag{S.7}$$

When the observable mobility data is reliable and works as an adequate representative of the social distancing measure, as described in our model, this direct estimate of $\alpha$ is likely to be close to the crude slope estimate of the regression problem described earlier.

#### S4. Doubling times and rates

With $C_t$ denoting the total confirmed cases per million up to time $t$, the *doubling time* $t_d(t)$ at time $t$ is the length of time necessary for the cumulative cases to double. In other words, $t_d(t)$ is defined explicitly by the relation

$$\frac{C_{(t+t_d(t))}}{C_t} = 2. \tag{S.8}$$

By a first order Taylor series expansion, a linear approximation of the numerator in (S.8) is

$$C_{(t+t_d(t))} = C_t + t_d(t)\frac{d}{dt}C_t. \tag{S.9}$$

(S.8) and (S.9) together implies

$$t_d(t) = \frac{C_t}{\frac{d}{dt}C_t} = \frac{1}{\frac{d}{dt}\log(C_t)}$$

The *doubling rate* $\tilde{\xi}(t)$ is defined as

$$\tilde{\xi}(t) = \frac{1}{t_d(t)} = \frac{d}{dt}\log(C_t). \tag{S.10}$$

The doubling rate quantifies the rate of spread, with lower doubling rates corresponding to longer doubling times and signify a better containment of the disease.

#### S5. Residual bootstrap procedure for inference

The estimated parameters can be used to construct confidence intervals for the relevant epidemiological quantities. To this end, we describe the steps for a residual bootstrap procedure in this context in the Algorithm 1

**Algorithm 1:** Residual Bootstrap Implementation

**Input:** $C_t$, $R_t$, $Q_t$,$H_t$, $D_t$, level = 0.95, Time Window

**Output:** 95% C.I. for $\boldsymbol{\beta}(t)$, $\boldsymbol{\zeta}$ and $\alpha$.

1 Set the initial values for $C_t$, $R_t$, $Q_t$,$H_t$, $D_t$ same as the observed quantity at the beginning of the pandemic.

2 Compute the estimated trajectories iteratively through the course of time.

$$\widehat{\Delta C}_t = (\widehat{\theta}(t) + \hat{\gamma})\widehat{A}_t; \quad \widehat{\Delta Q}_t = \widehat{\theta}(t)\widehat{A}_t - (\hat{\gamma} + \hat{\rho}_A)\hat{Q}_t.$$

$$\widehat{\Delta H}_t = \hat{\gamma}(\widehat{A}_t + \hat{Q}_t) - \widehat{\rho_H}(t)\hat{H}_t - \widehat{\delta}(t)\hat{H}_t; \quad \widehat{\Delta R^H}_t = \widehat{\rho_H}(t)\hat{H}_t; \quad \widehat{\Delta R^Q}_t = \hat{\rho}_A\hat{Q}_t$$

$$\widehat{\Delta R^A}_t = \hat{\rho}_A\widehat{A_t}; \quad \widehat{\Delta R}_t = \widehat{\Delta R^H}_t + \widehat{\Delta R^Q}_t + \widehat{\Delta R^A}_t; \quad \widehat{\Delta D}_t = \widehat{\delta}(t)\hat{H}_t.$$

3 Construct residuals as the difference between the observed values and the estimated numbers for each of these five states separately. For State $X$, $X \in \{C, D, R, H, Q\}$, the residual is computed as

$$e_{X_t} = X_t - \hat{X}_t.$$

4 Compute the mean adjusted residuals as

$$\tilde{e}_{X_t} = e_{X_t} - \overline{e_{X_t}}.$$

5 Resample from the mean adjusted residuals with replacement to construct $\tilde{e^*}_{X_t}$.

6 Construct a new bootstrap sample trajectory as,

$$X^{*(b)} = \hat{X}_t + \tilde{e^*}_{X_t} + \overline{e_{X_t}}.$$

7 Fit the model in Section 5 based on the new bootstrap samples and estimate the epidemiological quantities. For the $b$-th bootstrap sample we obtain the bootstrap estimates $\widehat{\boldsymbol{\beta}}^{*b}(t)$, $\widehat{\boldsymbol{\zeta}}^{*b}$ and $\widehat{\alpha}^{*b}$.

8 Repeat *Steps 5 to 7* for large number ($B$) of times.

9 Use percentile bootstrap procedure to find the confidence intervals with level 0.95 (pointwise C.I. for the time-dependent parameters respectively) based on the $B$ bootstrap estimate.

#### S6. Aggregation Simulation study based on the Poisson Model

In this section we present results of a simulation study in order to partially justify the proposed estimation procedure. We also use a parametric bootstrap approach to construct pointwise confidence intervals for the estimated rate parameters and other epidemiological quantities. We also compare these confidence intervals with those obtained from the residual bootstrap procedure.

S6.1. **Data Generation and Aggregation.** Given the values of the parameters, the simulated trajectories are modeled as linear combinations of sub-Poisson processes. To this end, we look at the individual drivers or branches that generate the dynamics at a granular level (see Figure 1 in Section 2.1)

$$\theta(t) = \phi(t)\frac{\Delta T_t}{H_t}, \tag{S.11}$$

$$P_{tA} \sim Poisson(\theta(t)A_t), \quad P_{gA} \sim Poisson(\gamma A_t), \tag{S.12}$$

$$P_{rA} \sim Poisson(\rho_A A_t), \quad P_{aA} \sim Poisson\left(\frac{S_t}{S_t + R_t + A_t}\right)\alpha\kappa^2(t)A_t, \tag{S.13}$$

$$P_{gQ} \sim Poisson(\gamma Q_t), \quad P_{rQ} \sim Poisson(\rho_A Q_t), \tag{S.14}$$

$$P_{rH} \sim Poisson(\rho_H(t)H_t), \quad P_d \sim Poisson(\delta(t)H_t), \tag{S.15}$$

$$\Delta A_t = -(P_{tA} + P_{gA} + P_{rA}) + P_{aA}, \quad \Delta Q_t = P_{tA} - (P_{gQ} + P_{rQ}), \tag{S.16}$$

$$\Delta H_t = P_{gA} + P_{gQ} - (P_{rH} + P_d), \quad \Delta D_t = P_d, \quad \Delta C_t = P_{tA} + P_{gA}, \tag{S.17}$$

$$\Delta S_t = -\Delta A_t, \quad \Delta R_t^{reported} = \Delta R_t^Q + \Delta R_t^H = P_{rQ} + P_{rH}. \tag{S.18}$$

In the above simulation (S.11) – (S.18) are the Poisson process random variable versions of the population model (1) – (7) (see Section 2.1) Here, we assume $T_t, \kappa_t$ to be given. Also, in compliance with assumption **A5**, we take $\rho_Q(t) \equiv \rho_A$. Further, in (S.18) we use $\Delta R_t^{reported}$ instead of $\Delta R_t = \Delta R_t^A + \Delta R_t^Q + \Delta R_t^H$ since only $R_t^{reported} = R_t^Q + R_t^H$ can be observed or reported in practice. The values for $\Delta A_t, \Delta H_t, \Delta Q_t$ can be negative.

In the beginning of the pandemic the number of individuals in the symptomatic or quarantined states are quite low, introducing an inherent bias in the estimated trajectories. We use an aggregation procedure to increase the size of the simulated samples.

The general idea of the aggregation method is to make use of the fixed intrinsic parameters such as $\gamma$ and $\rho_A$ to generate 10 independent processes as 10 locations, for example counties or cities and consider the loss function to be the sum of the loss of these 10 independent processes. For the $i^{\text{th}}$ state or process, we have the loss metric $d_s^{(i)}$ as in (S.2) and the new locally weighted loss function conditional on $\boldsymbol{\zeta} = (\gamma, \rho_A)$ is constructed as

$$\widetilde{L}_t^h(\boldsymbol{\beta}(t)|\boldsymbol{\zeta}) = \sum_{i=1}^{10} \sum_s \frac{1}{h} K\left(\frac{t-s}{h}\right) d_s^{(i)}(\boldsymbol{\beta}(t)|\boldsymbol{\zeta}). \tag{S.19}$$

The time varying parameters are estimated as the minimiser of (S.19), while the intrinsic parameters are found as the optimizers of the corresponding profile loss function using a grid search algorithm.

S6.2. **Parametric Bootstrap Algorithm.** We now describe a parametric bootstrap method to construct pointwise confidence intervals for the estimated rate parameters and other relevant epidemiological quantities.

Step 1. Given the estimates of the parameters $\widehat{\boldsymbol{\beta}}(t)$, $\widehat{\boldsymbol{\zeta}}$ and $\widehat{\alpha}$ (assuming $\kappa_t$ is observed), simulate multiple copies of the data from a Poisson process with the dynamical states given by (1) – (7) in Section 2.1, by substituting the true parameter values with the estimates, over the same time domain, treating $T_t$ (number of tests) as given (since this is an intervention process that does not have a discernible generative mechanism).

Step 2. For the $k$-th sample, obtain the bootstrap estimates $\widehat{\boldsymbol{\beta}}^{*k}(t)$, $\widehat{\boldsymbol{\zeta}}^{*k}$ and $\widehat{\alpha}^{*k}$.

Step 3. Use percentile bootstrap procedure to find the confidence intervals (pointwise C.I. for the time-dependent parameters respectively).

S6.3. **Simulated Data and Results.** The simulation setting is studied for $T = 100$ days for 10 independent processes. The true parameters generating the processes are chosen as follows. The time-invariant parameters are: $\gamma = 0.03$, indicating that on a certain day 3 of 100 asymptomatic individuals may become symptomatic, $\rho_A = 0.06$, meaning daily 6 among 100 asymptomatic individuals may recover, and $\alpha = 0.2$ as the baseline infection rate, which suggests 20 of 100 susceptible individuals may be infected without social distancing The recovering rate from state $H_t$ is time-dependent: 0.03 for the first 20 days, 0.04 for the following 40 days, and 0.05 for the rest of the days. $\phi(t)$, testing efficiency, is a linear function of time $T$ with 0.00005 as a small positive slope and 0.001 as the intercept which reflects increasing testing efficiency. the intrinsic parameters and mobility index are fixed across all processes. The mobility index $\kappa_t$ is also a time-dependent parameter, ranging from 0.4 to 1.2 to describe the overall social distancing of people. $\delta(t)$ represents time-dependent death rate over 100 days. It is set as 0.007 during the first 50 days while 0.005 during the following 50 days. That is, 7 out of $1,000$ hospitalized individuals may die during the first half of the period while 5 out of $1,000$ hospitalized ones may die during the second half. Each process will be assigned to different numbers of test cases.

S6.3.1. *Parametric Bootstrap.* We first conduct a grid search for $\gamma$ and $\rho_A$ to estimate the optimal parameters minimizing the loss function (see (20) in Section 5.1) This yields

$$\hat{\gamma} = 0.027, \hat{\rho}_A = 0.06.$$

We then estimate the other time-dynamic parameters by using the values of $\hat{\gamma}, \hat{\rho}_A$.

Using the parametric bootstrap procedure (Section S6), we compute the bootstrap sampling distributions and confidence regions for various epidemiological parameters and compare them with the true values of the underlying parameters (See Figure 2).

Estimation of the other releavant quantities, such as $A_t$ - the number of infected but asymptomatic people at time $t$ and $\Delta C_t$ - the number of confirmed cases upto time $t$, are

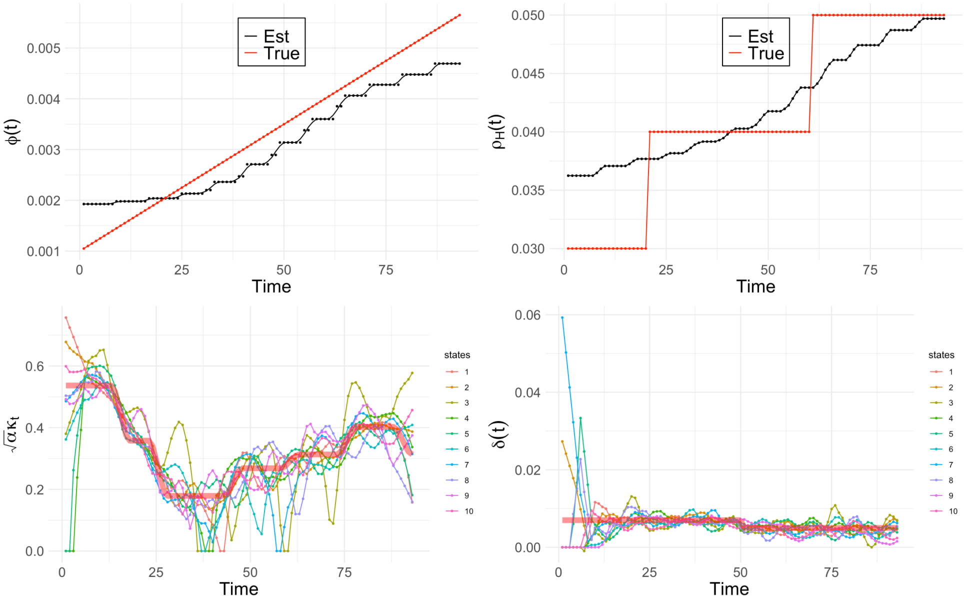


FIGURE 1. From left to right : plots of estimations for $\phi(t), \rho_H(t)$, $\sqrt{\alpha}\kappa_t$, and $\delta(t)$; the wide red lines denote the true $\sqrt{\alpha}\kappa_t$ and $\delta(t)$

also performed, and point-wise confidence bands provided using a parametric bootstrap mechanism (See Figure 3 and 4).

We observe that the estimates of $\gamma$ and $\rho_A$ are very close to our true values. Figure 1 reports the estimates against the true parameters. $\hat{\phi}(t)$ and $\widehat{\rho_H}(t)$ are quite close to the true ones, with a bias smaller than 0.01 and capturing the overall pattern of change. The estimates of $\sqrt{\alpha}\kappa_t$ and $\delta(t)$ are varying across the 10 processes, however, the overall trends are captured well.

Under the same true parameter setting, we now generate 10 independent copies of the aggregation processes and repeat the estimation procedure 1000 times, which yields a set of estimates for the parameters and hence the bootstrap sampling distributions respectively. We notice that, the 95% confidence interval constructed based on the bootstrap sampling distribution contains the true values of $\gamma = 0.03$ and $\rho_A = 0.06$. It is also found that the true $\phi(t)$ lies within the corresponding confidence band uniformly over time. The true $\rho_H(t)$ intersects partially with the respective confidence region, however, the discrepancy,

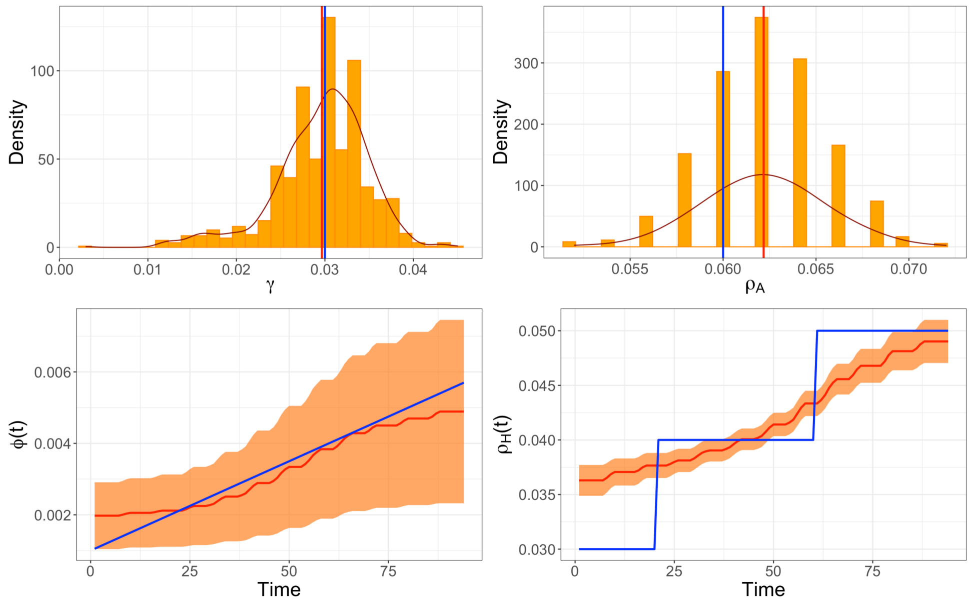


FIGURE 2. From left to right: the top row displays the sampling distribution of $\gamma$ and $\rho_A$ - the bootstrap sample mean and the true parameter values are depicted in the red line and the darkgreen lines respectively. The bottom row show the 95% point-wise confidence intervals for $\phi(t)$ and $\rho_H(t)$ based on 1000 bootstrap samples- the bootstrap sample means are shown in red while the blue curves are the true parameters.

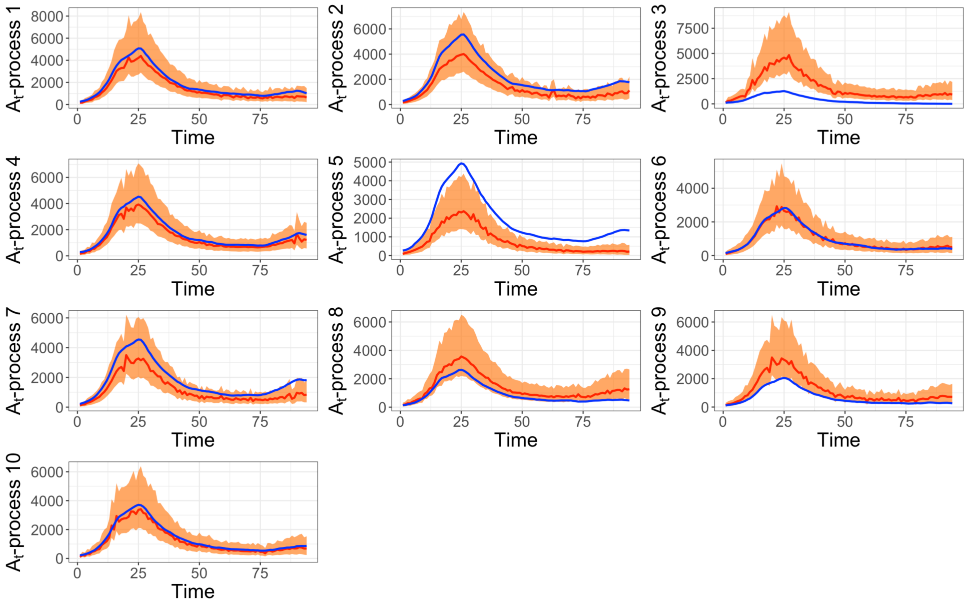


FIGURE 3. Point-wise Confidence Interval for $A_t$ for all 10 independent processes; point-wise mean is in red and the true data is in blue.

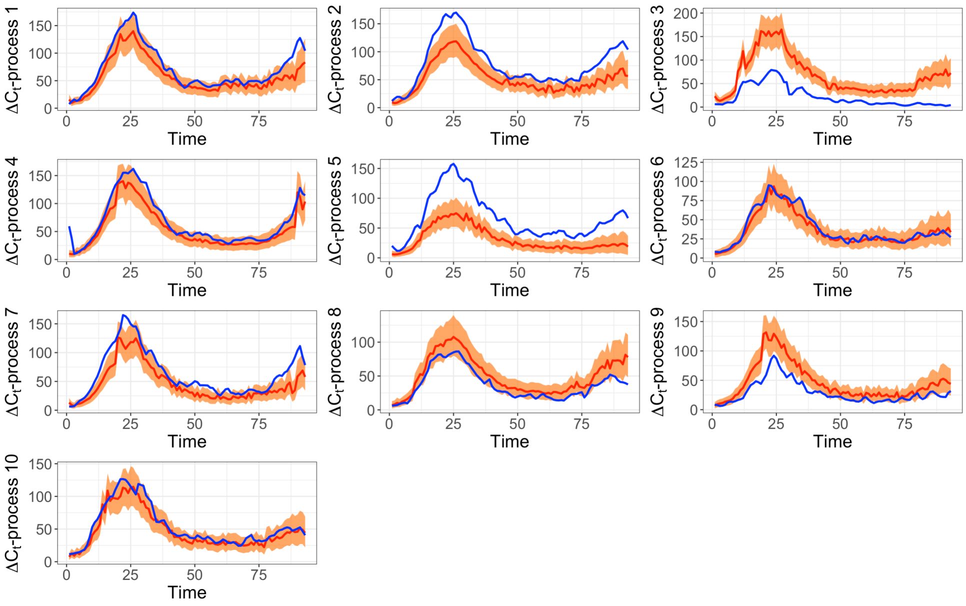

FIGURE 4. Pointwise Confidence Interval for $\Delta C_t$; point-wise mean is in red and the true data is in blue.

where it lies outside, is as small as 0.005. The 95% confidence intervals for $A_t$ and $\Delta C_t$ with their true generated trajectories are shown in Figure 3 and 4. Here again the estimations from the simulated data are falling or very close to the confidence regions.

We can conclude, based on the simulation results, that the Poisson process model is able to describe the dynamics pretty well, as long as we have enough data and valid information. However, due to the limited sample size and possible lack of valid information, the data we encounter in real life are noisier, resulting in biased estimates if the Poisson process model is applied directly.

S6.3.2. *Residual Bootstrap.* We apply a residual bootstrap strategy to the same simulation setting (as described in Section S6.3) and compare the performances of the two types of bootstrap inference. We observed that, the sampling distribution of $\gamma$ and $\rho_A$ for the residual bootstrap (Figure 5) have wider variations than those obtained by the parametric bootstrap method (Figure 2). A similar occurrence is noted for $\phi(t)$ while comparing the two confidence regions obtained by the two different bootstrap strategies. However, the confidence regions for $\rho_H(t)$ are roughly the same. In addition, these two strategies, parametric and residual bootstrap, show difference in $A_t$ and $C_t$ (Figure 6 and 7). It turns out the residual bootstrap implies much wider confidence regions of $A_t$ but narrower ones of $\Delta C_t$, compared to those by the parametric bootstrap. Despite these differences, both the methods indicate a fair performance when it comes to inference, noticing that the true values for the parameters mostly lie within or very close to the confidence regions. In general, the parametric bootstrap gives a narrower confidence band, but can only be justified when there is ample data to support the underlying assumption of the Poisson process mechanism. The residual bootstrap, being nonparametric on the other hand, is data-driven, robust, and hence more adoptable to the current data analysis paradigm, despite the fact that the confidence regions provided are wider.

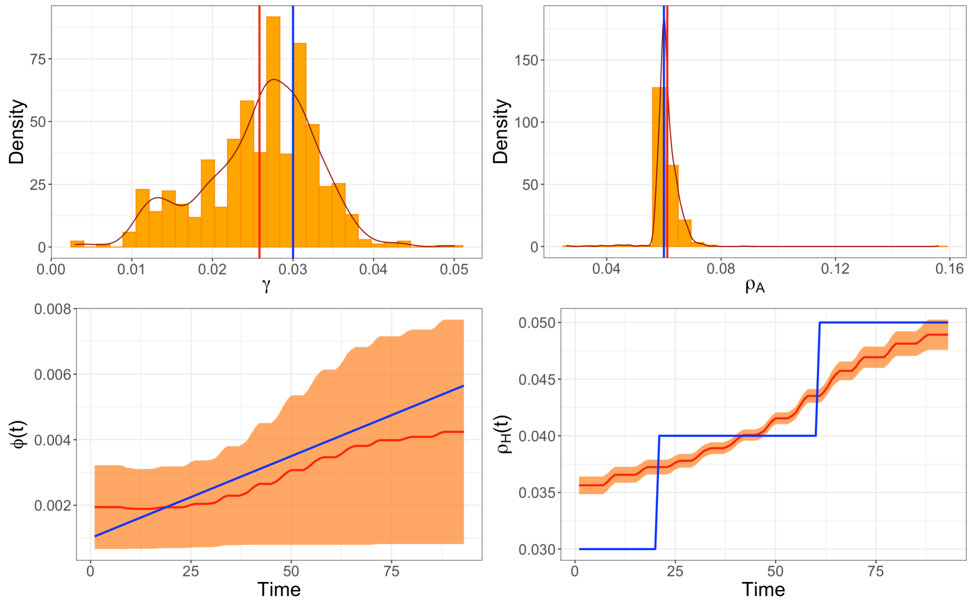


FIGURE 5. From left to right: the top row displays the sampling distribution of $\gamma$ and $\rho_A$ - the bootstrap sample mean and the true parameter values are depicted in the red line and the darkgreen lines respectively. The bottom row show the 95% point-wise confidence intervals for $\phi(t)$ and $\rho_H(t)$ based on 1000 bootstrap samples- the bootstrap sample means are shown in red while the blue curves are the true parameters.

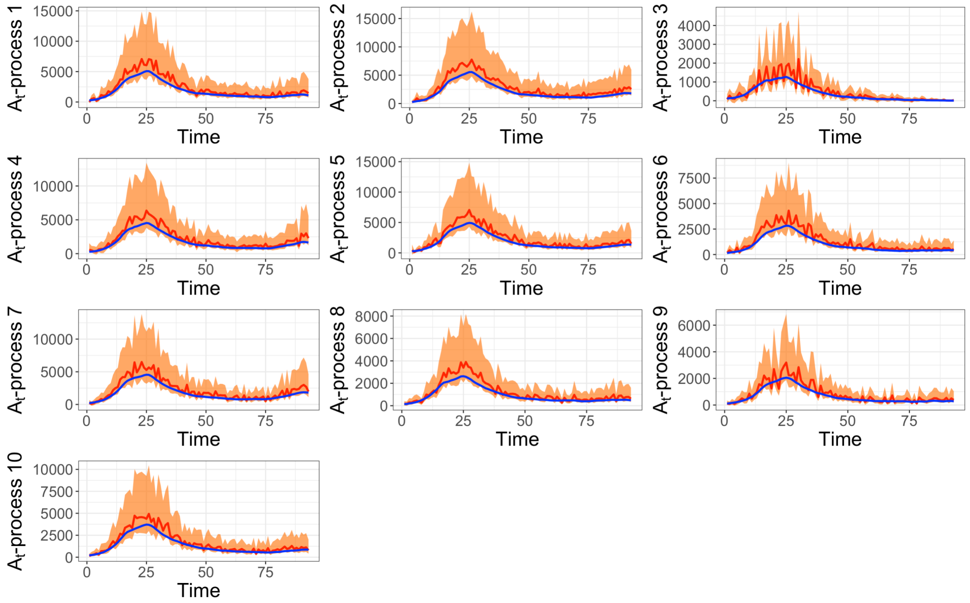

FIGURE 6. Point-wise Confidence Interval for $A_t$ for all 10 independent processes; point-wise mean is in red and the true data is in blue.

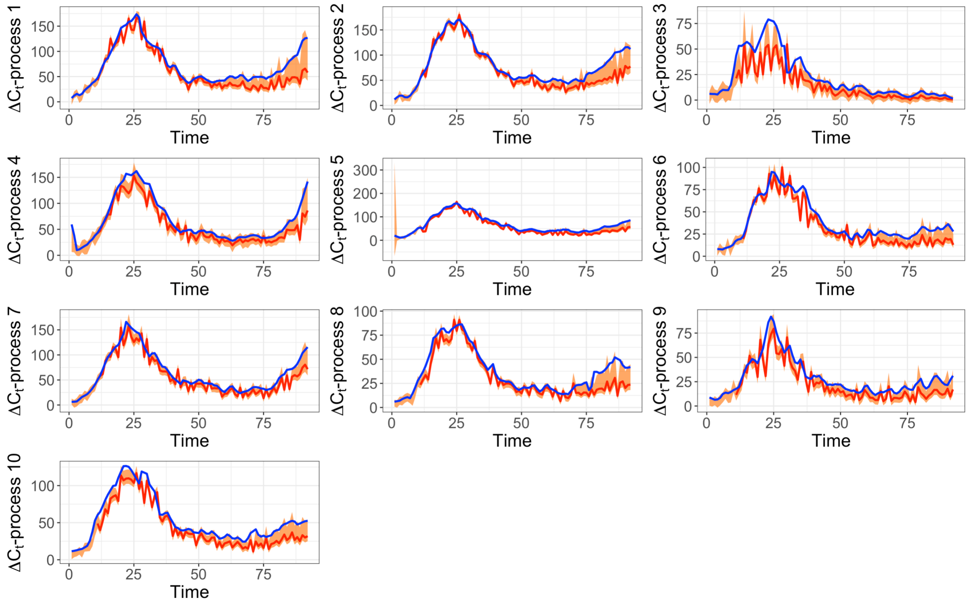

FIGURE 7. Pointwise Confidence Interval for $\Delta C_t$; point-wise mean is in red and the true data is in blue.

#### S7. Additional results for a select few states of the USA

The table comparing the estimates for the relevant parameters for some chosen states of the USA (Table 2 in Section 3.2.2) is extended to include the estimates of $\alpha$ - the basic infection rate without any social distancing measure put in place, and $\phi(t)$ - the time-varying parameter testing efficiency. To this end, the estimate of $\alpha$ is found from (S.7), as described in Supplement Section S2, using the google mobility data as a surrogate for $\kappa(t)$. The mean estimates for the time varying parameters $\delta(t)$, $\rho_H(t)$, $\theta(t)$, and $\phi(t)$ are shown (see Table 1).

| | $\hat{\gamma}$ | $\hat{\rho}_A$ | $\hat{\alpha}$ | $\widehat{\delta(t)}$ | $\widehat{\rho_H(t)}$ | $\widehat{\theta(t)}$ | $\widehat{\phi(t)}$ |
|---|---|---|---|---|---|---|---|
| Arizona | 0.0003 | 0.002 | 0.1509 | 0.0208 | 0.0023 | 0.0887 | 0.0079 |
| Arkansas | 0.0029 | 0.094 | 0.1775 | 0.0249 | 0.0975 | 0.0809 | 0.0038 |
| Delaware | 0.0017 | 0.008 | 0.1519 | 0.0159 | 0.0093 | 0.1076 | 0.0037 |
| Idaho | 0.0009 | 0.010 | 0.0357 | 0.0230 | 0.0138 | 0.0289 | 0.0019 |
| Iowa | 0.0011 | 0.032 | 0.0682 | 0.0263 | 0.0372 | 0.0478 | 0.0033 |
| Minnesota | 0.0023 | 0.128 | 0.2410 | 0.0315 | 0.0654 | 0.0899 | 0.0034 |
| Nebraska | 0.0011 | 0.020 | 0.0557 | 0.0141 | 0.0266 | 0.0394 | 0.0035 |
| Ohio | 0.0023 | 0.048 | 0.1027 | 0.0180 | 0.0532 | 0.0625 | 0.0024 |
| Oklahoma | 0.0037 | 0.084 | 0.1372 | 0.0122 | 0.1029 | 0.0494 | 0.0033 |
| Pennsylvania | 0.0013 | 0.026 | 0.0818 | 0.0293 | 0.0372 | 0.0535 | 0.0033 |
| South Dakota | 0.0021 | 0.058 | 0.0561 | 0.0190 | 0.0922 | 0.0262 | 0.0038 |
| Tennessee | 0.0059 | 0.064 | 0.1907 | 0.0158 | 0.0413 | 0.1206 | 0.0076 |
| Texas | 0.0019 | 0.036 | 0.0986 | 0.0207 | 0.0341 | 0.0212 | 0.0013 |
| Utah | 0.0011 | 0.040 | 0.1692 | 0.0144 | 0.0252 | 0.1434 | 0.0061 |
| Wisconsin | 0.0017 | 0.068 | 0.0926 | 0.0217 | 0.0707 | 0.0477 | 0.0026 |

Table 1. Mean estimated parameters for different states in the USA.

We now present the point estimates, as well as the residual bootstrap results, for the relevant parameters and epidemiological markers pertaining to a select few US states. The results can be interpreted in a similar way as for the case of Utah in Section 3.2.1.

### Arizona

| | Estimate | 95% Confidence Interval | Mean | s.d. |
|---|---|---|---|---|
| $\gamma$ | 0.0003 | [0.0019, 0.0032] | 0.0018 | 0.0031 |
| $\rho_A$ | 0.0020 | [0.0200, 0.0320] | 0.0275 | 0.0011 |
| $\alpha$ | 0.1509 | [0.1514, 0.1684] | 0.1580 | 0.0044 |

TABLE 2. Confidence intervals, mean and standard deviations for the time-invariant parameters, computed based on 1000 bootstrap samples using residual bootstrap approach for *Arizona*.

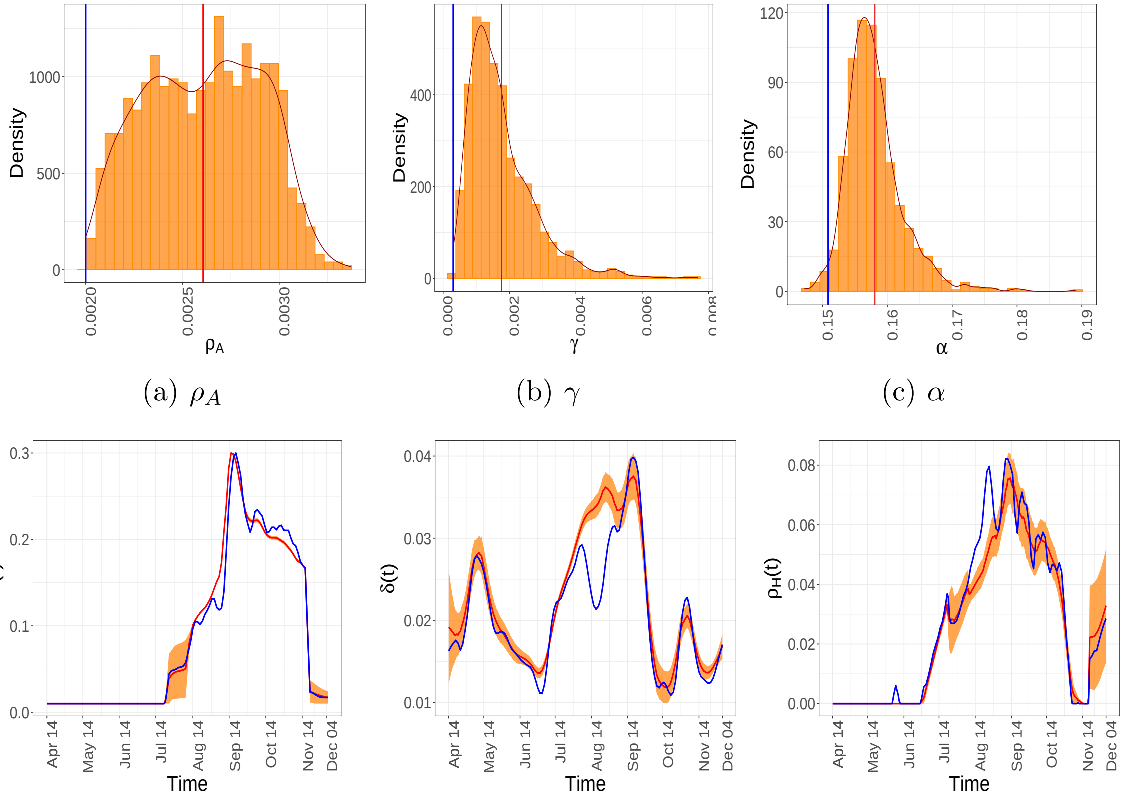


(a) $\rho_A$ (b) $\gamma$ (c) $\alpha$

(d) $\theta(t)$ (e) $\delta(t)$ (f) $\rho_H(t)$

FIGURE 8. Estimates and residual bootstrap based confidence intervals for time invariant and time-varying parameters for the state of *Arizona*. The estimate from the data is in blue. The 95% confidence band is in yellow and the mean of the bootstrap estimates are presented in red.

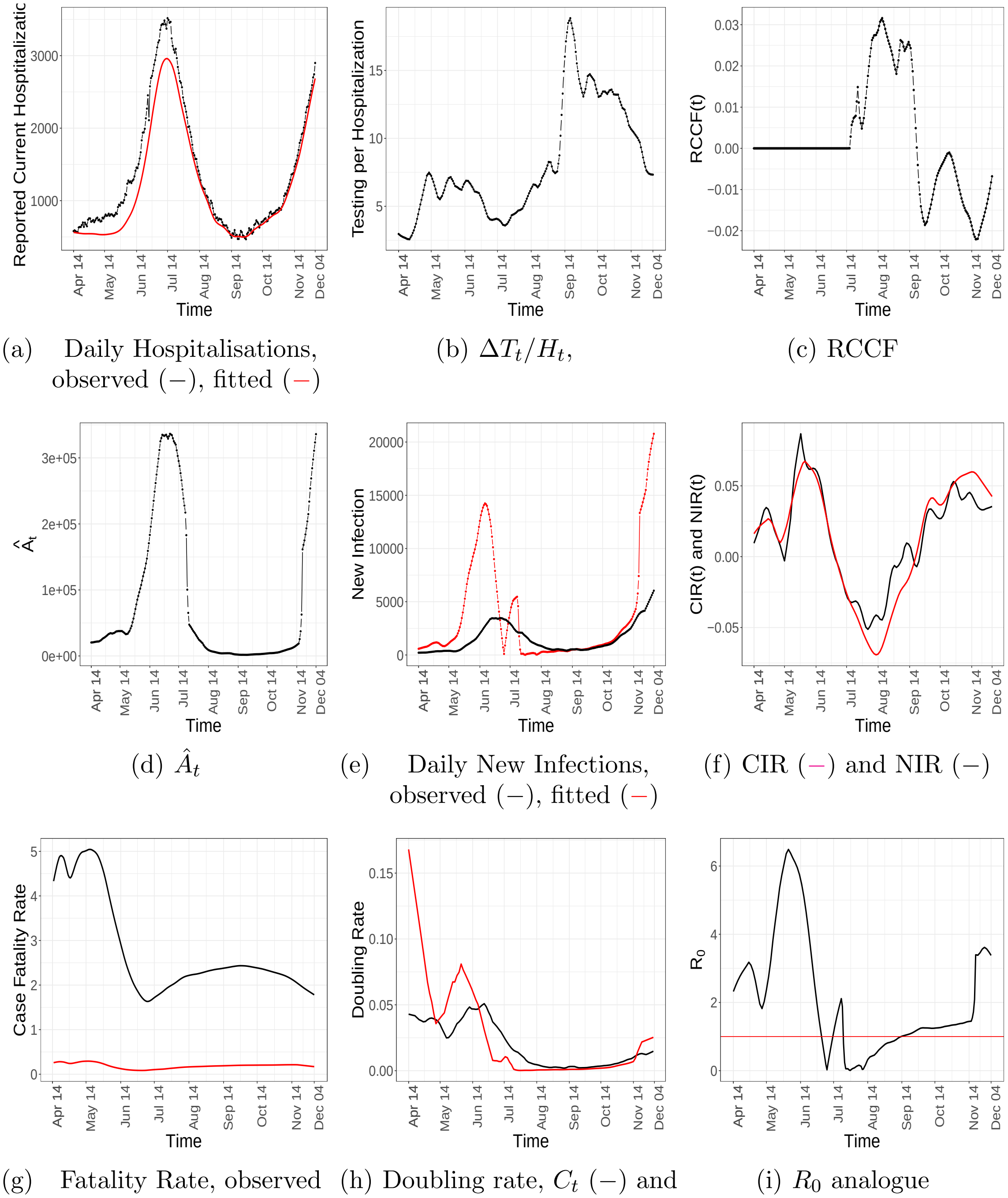


(a) Daily Hospitalisations, observed (–), fitted (–)

(b) $\Delta T_t/H_t$,

(c) RCCF

(d) $\hat{A}_t$

(e) Daily New Infections, observed (–), fitted (–)

(f) CIR (–) and NIR (–)

(g) Fatality Rate, observed (–), actual (–)

(h) Doubling rate, $C_t$ (–) and $CNI$ (–)

(i) $R_0$ analogue

FIGURE 9. Temporal patterns of some components and epidemiological markers for *Arizona*.

### Arkansas

| | Estimate | 95% Confidence Interval | Mean | s.d. |
|---|---|---|---|---|
| $\gamma$ | 0.0029 | [0.0023, 0.0057] | 0.0037 | 0.0009 |
| $\rho_A$ | 0.0940 | [0.0760, 0.1020] | 0.0902 | 0.0007 |
| $\alpha$ | 0.1775 | [0.1561, 0.2100] | 0.1870 | 0.0142 |

Table 3. Confidence intervals, mean and standard deviations for the time-invariant parameters, computed based on 1000 bootstrap samples using residual bootstrap approach for *Arkansas*.

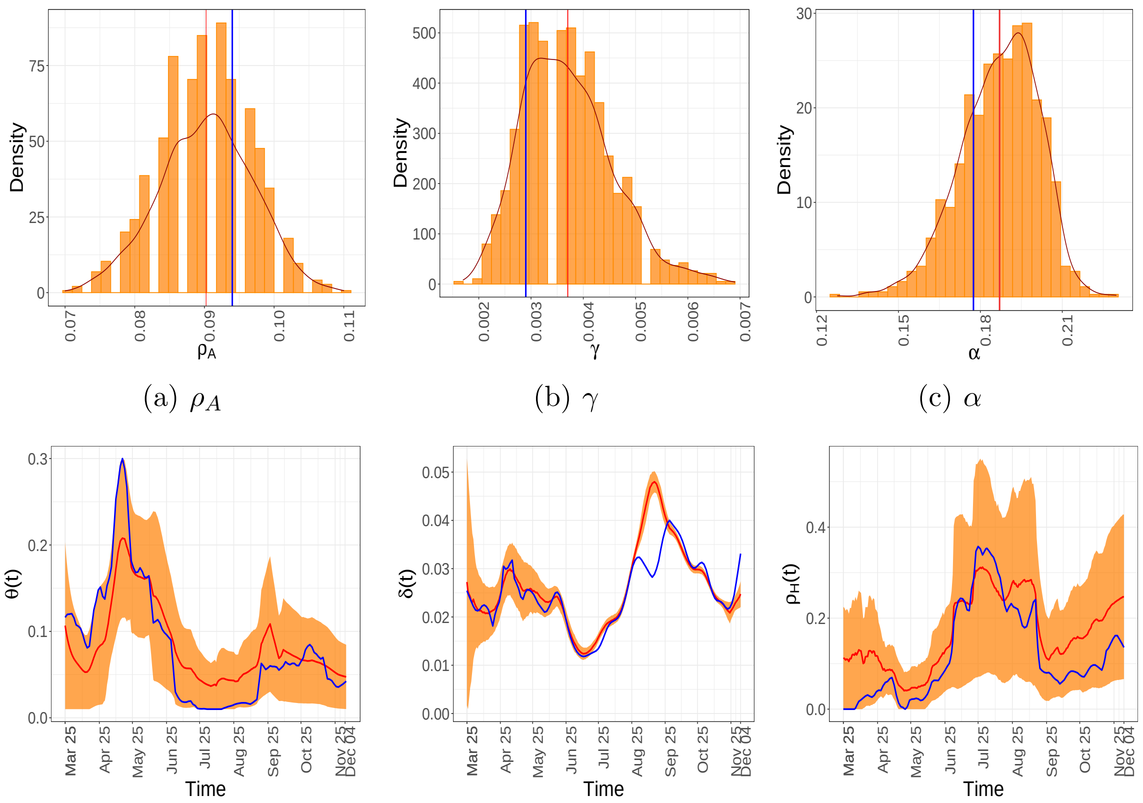


(a) $\rho_A$ (b) $\gamma$ (c) $\alpha$

(d) $\theta(t)$ (e) $\delta(t)$ (f) $\rho_H(t)$

Figure 10. Estimates and residual bootstrap based confidence intervals for time invariant and time-varying parameters for the state of *Arkansas*. The estimate from the data is in blue. The 95% confidence band is in yellow and the mean of the bootstrap estimates are presented in red.

The point estimates and bootstrap confidence for the relevant parameters are presented below. The mortality and hospitalization rate in Arkansas is high.

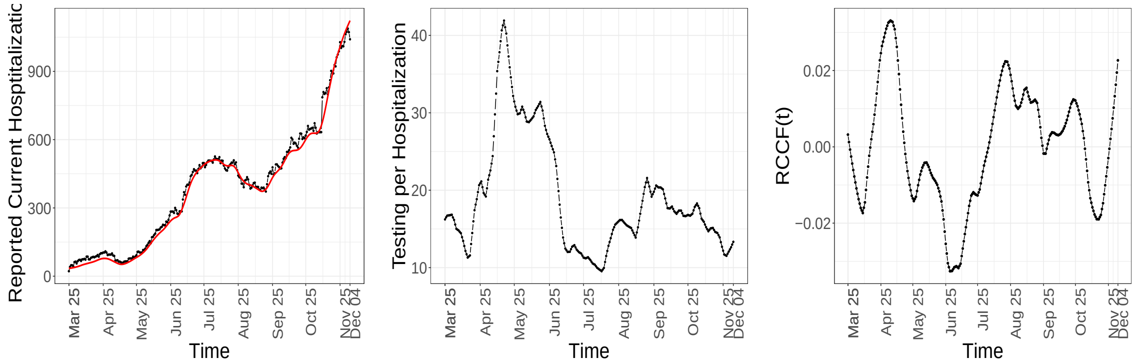


(a) Daily Hospitalisations, observed (−), fitted (−)
(b) $\Delta T_t/H_t$,
(c) RCCF

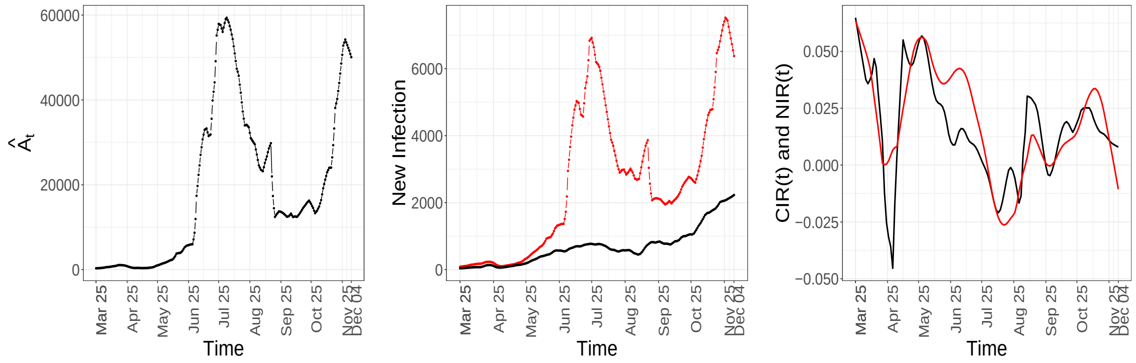


(d) $\hat{A}_t$
(e) Daily New Infections, observed (−), fitted (−)
(f) CIR (−) and NIR (−)

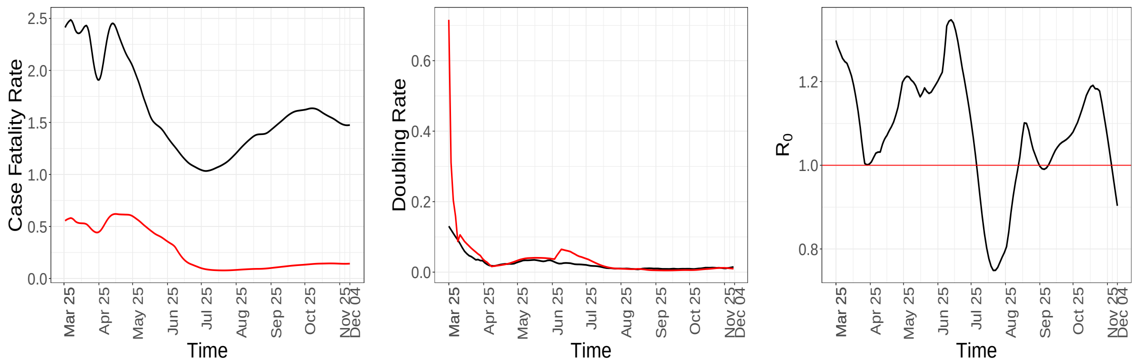


(g) Fatality Rate, observed (−), actual (−)
(h) Doubling rate, $C_t$ (−) and $CNI$ (−)
(i) $R_0$ analogue

FIGURE 11. Temporal patterns of some components and epidemiological markers for *Arkansas*.

### Delaware

| | Estimate | 95% Confidence Interval | Mean | s.d. |
|---|---|---|---|---|
| $\gamma$ | 0.0017 | [0.0017, 0.0021] | 0.0019 | 0.0009 |
| $\rho_A$ | 0.0080 | [0.0080, 0.0087] | 0.0081 | 0.0001 |
| $\alpha$ | 0.1519 | [0.0921, 0.1302] | 0.1140 | 0.0093 |

TABLE 4. Confidence intervals, mean and standard deviations for the time-invariant parameters, computed based on 1000 bootstrap samples using residual bootstrap approach for *Delaware*.

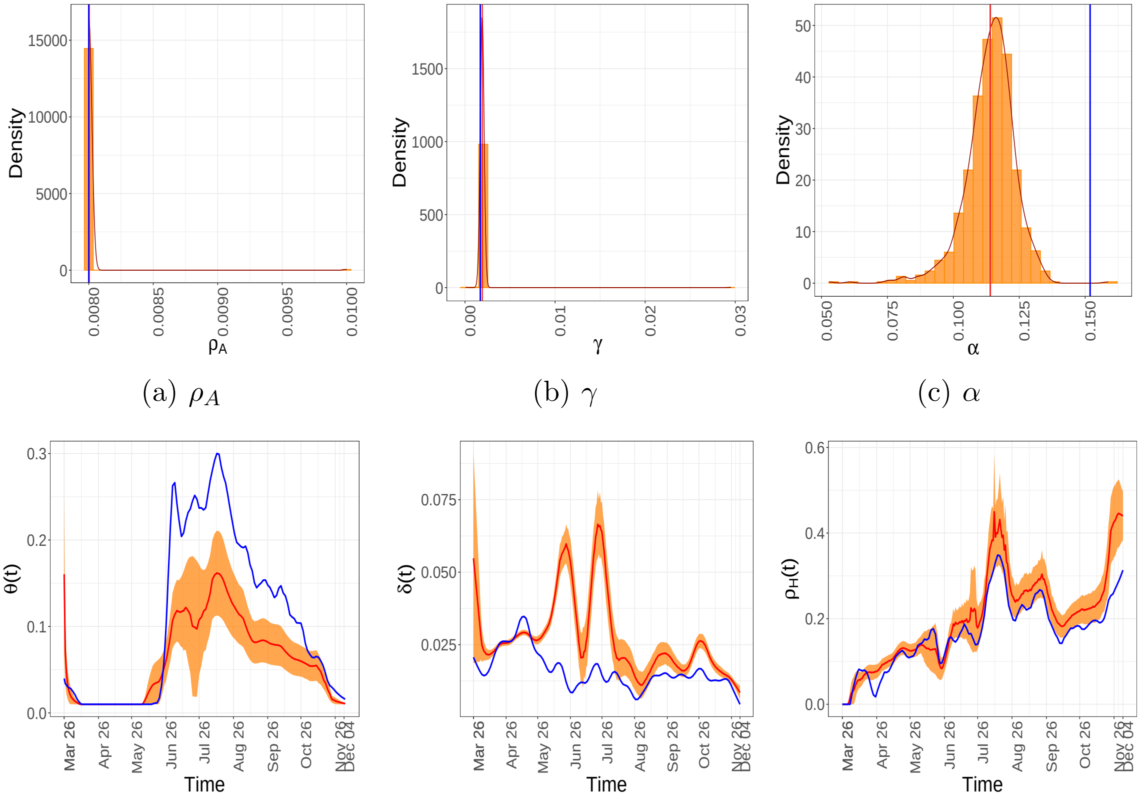


(a) $\rho_A$ (b) $\gamma$ (c) $\alpha$

(d) $\theta(t)$ (e) $\delta(t)$ (f) $\rho_H(t)$

FIGURE 12. Estimates and residual bootstrap based confidence intervals for time invariant and time-varying parameters for the state of *Delaware*. The estimate from the data is in blue. The 95% confidence band is in yellow and the mean of the bootstrap estimates are presented in red.

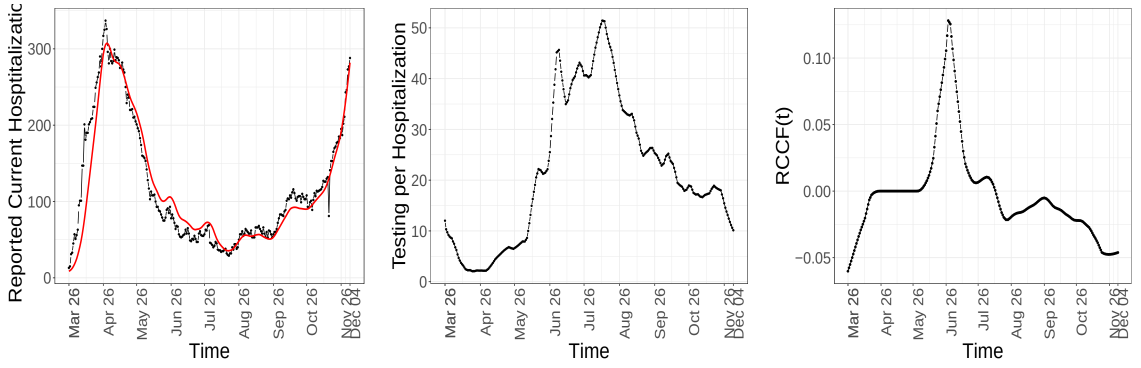


(a) Daily Hospitalisations, observed (–), fitted (–)

(b) $\Delta T_t/H_t$,

(c) RCCF

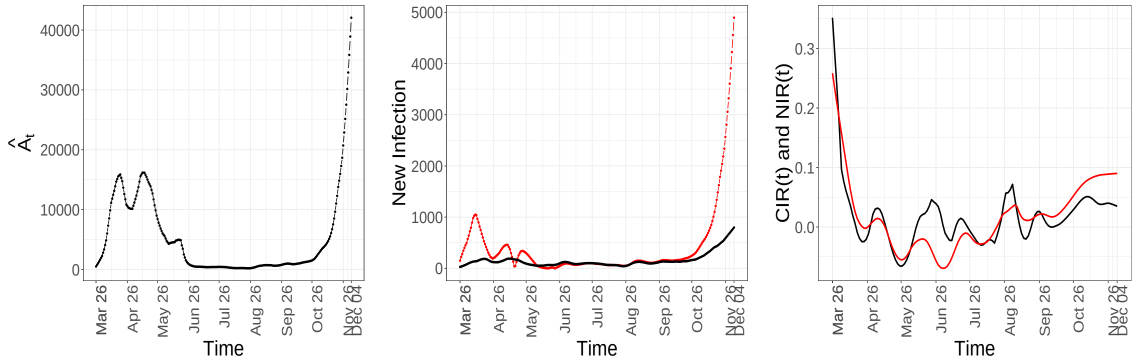


(d) $\hat{A}_t$

(e) Daily New Infections, observed (–), fitted (–)

(f) CIR (–) and NIR (–)

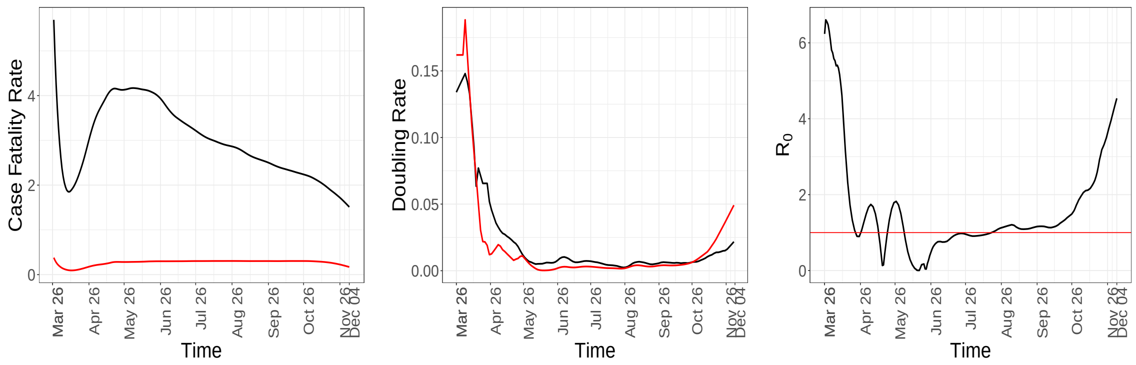


(g) Fatality Rate, observed (–), actual (–)

(h) Doubling rate, $C_t$ (–) and $CNI$ (–)

(i) $R_0$ analogue

FIGURE 13. Temporal patterns of some components and epidemiological markers for *Delaware*.

### Idaho

| | Estimate | 95% Confidence Interval | Mean | s.d. |
|---|---|---|---|---|
| $\gamma$ | 0.0009 | [0.0007, 0.0017] | 0.0011 | 0.0003 |
| $\rho_A$ | 0.0100 | [0.0080, 0.0100] | 0.0097 | 0.0007 |
| $\alpha$ | 0.0357 | [0.0385, 0.0639] | 0.0509 | 0.0216 |

TABLE 5. Confidence intervals, mean and standard deviations for the time-invariant parameters, computed based on 1000 bootstrap samples using residual bootstrap approach for *Idaho*.

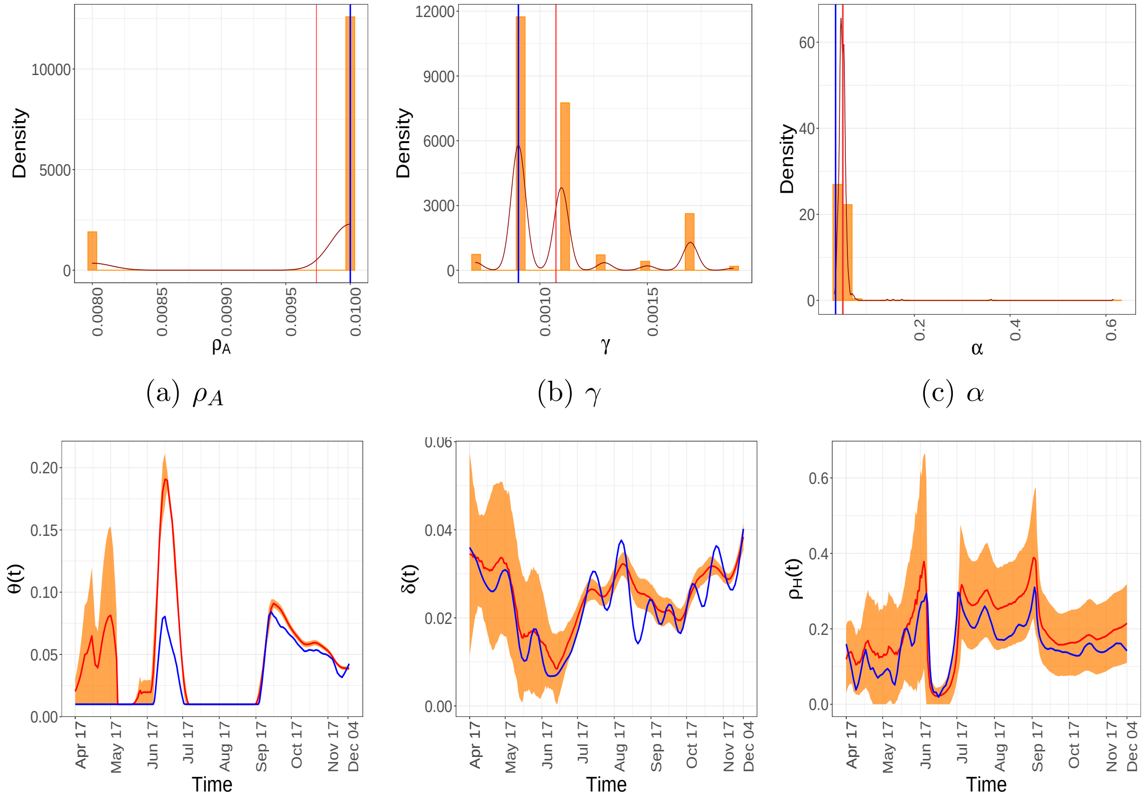


(a) $\rho_A$ (b) $\gamma$ (c) $\alpha$

(d) $\theta(t)$ (e) $\delta(t)$ (f) $\rho_H(t)$

FIGURE 14. Estimates and residual bootstrap based confidence intervals for time invariant and time-varying parameters for the state of *Idaho*. The estimate from the data is in blue. The 95% confidence band is in yellow and the mean of the bootstrap estimates are presented in red.

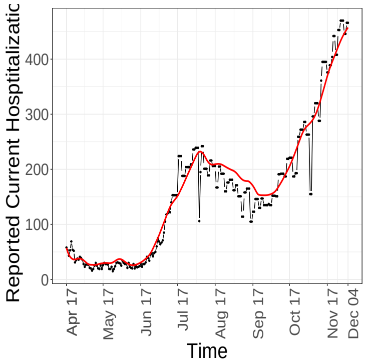


(a) Daily Hospitalisations, observed (–), fitted (–)

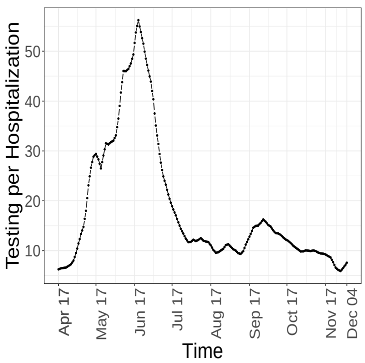


(b) $\Delta T_t / H_t$,

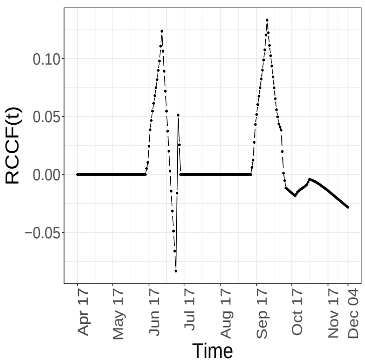


(c) RCCF

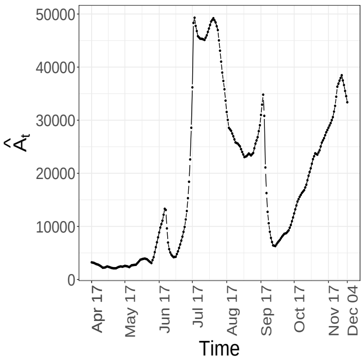


(d) $\hat{A}_t$

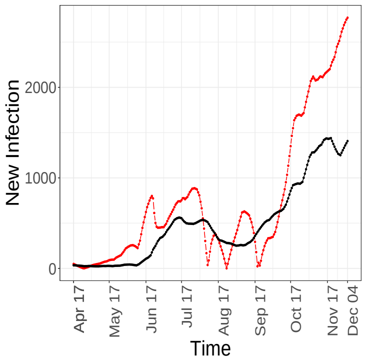


(e) Daily New Infections, observed (–), fitted (–)

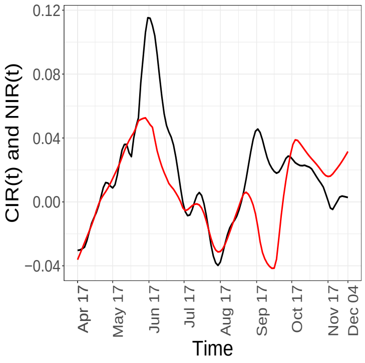


(f) CIR (–) and NIR (–)

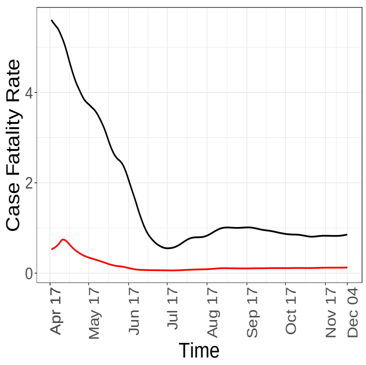


(g) Fatality Rate, observed (–), actual (–)

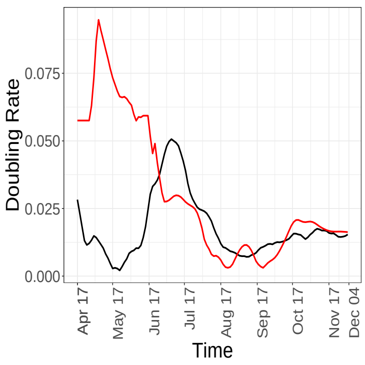


(h) Doubling rate, $C_t$ (–) and $CNI$ (–)

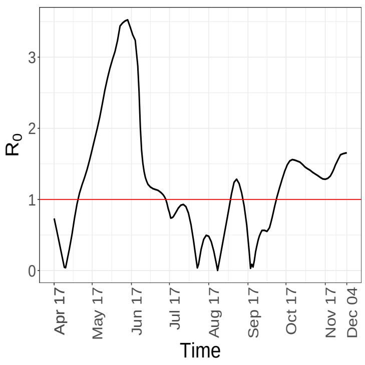


(i) $R_0$ analogue

FIGURE 15. Temporal patterns of some components and epidemiological markers for *Idaho*.

### Iowa

| | Estimate | 95% Confidence Interval | Mean | s.d. |
|---|---|---|---|---|
| $\gamma$ | 0.0011 | [0.0015, 0.0027] | 0.0021 | 0.0003 |
| $\rho_A$ | 0.0320 | [0.0180, 0.0260] | 0.0228 | 0.0021 |
| $\alpha$ | 0.0682 | [0.0407, 0.0798] | 0.0584 | 0.0101 |

TABLE 6. Confidence intervals, mean and standard deviations for the time-invariant parameters, computed based on 1000 bootstrap samples using residual bootstrap approach for *Iowa*.

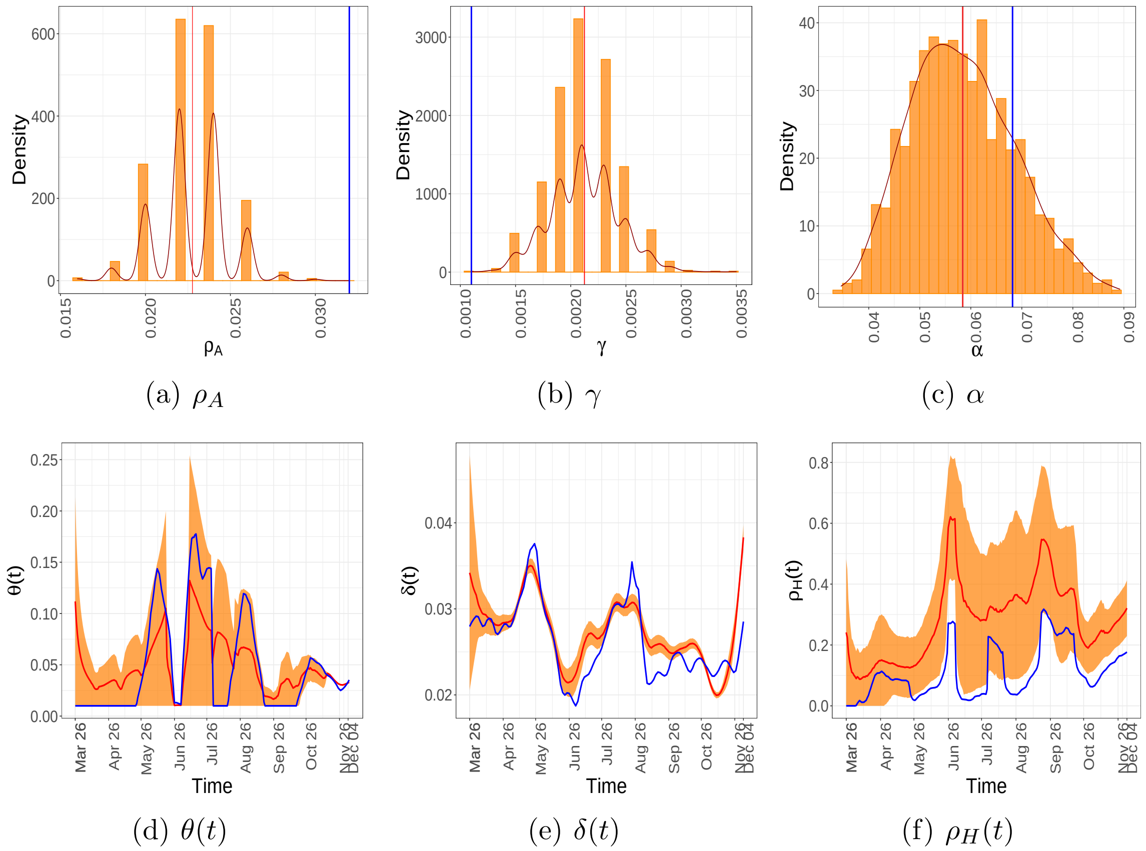


(a) $\rho_A$ (b) $\gamma$ (c) $\alpha$

(d) $\theta(t)$ (e) $\delta(t)$ (f) $\rho_H(t)$

FIGURE 16. Estimates and residual bootstrap based confidence intervals for time invariant and time-varying parameters for the state of *Iowa*. The estimate from the data is in blue. The 95% confidence band is in yellow and the mean of the bootstrap estimates are presented in red.

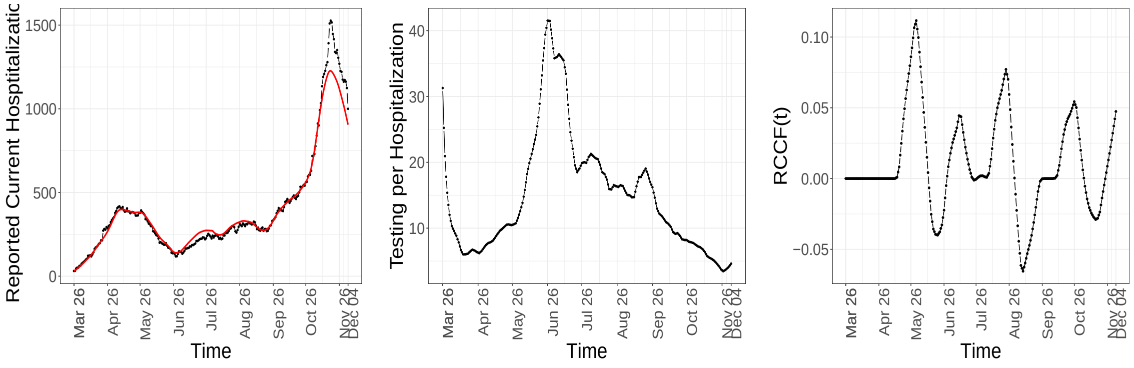


(a) Daily Hospitalisations, observed (–), fitted (–)

(b) $\Delta T_t / H_t$,

(c) RCCF

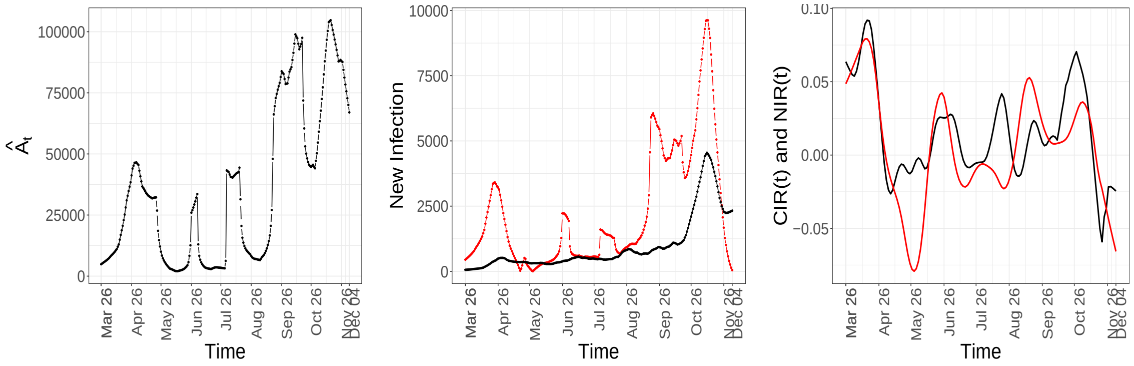


(d) $\hat{A}_t$

(e) Daily New Infections, observed (–), fitted (–)

(f) CIR (–) and NIR (–)

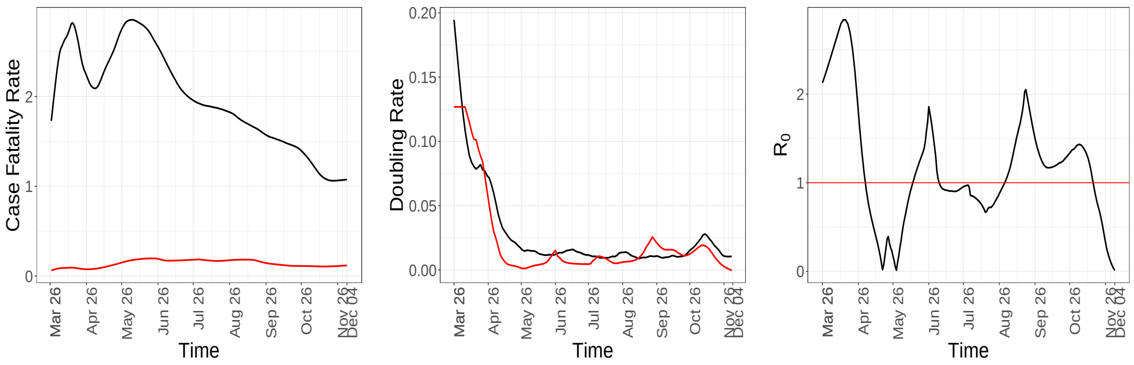


(g) Fatality Rate, observed (–), actual (–)

(h) Doubling rate, $C_t$ (–) and $CNI$ (–)

(i) $R_0$ analogue

FIGURE 17. Temporal patterns of some components and epidemiological markers for *Iowa*.

### Minnesota

| | Estimate | 95% Confidence Interval | Mean | s.d. |
|---|---|---|---|---|
| $\gamma$ | 0.0023 | [0.0021, 0.0183] | 0.0048 | 0.0043 |
| $\rho_A$ | 0.1280 | [0.0560, 0.0200] | 0.1142 | 0.0249 |
| $\alpha$ | 0.2410 | [0.1103, 0.3469] | 0.2105 | 0.0460 |

TABLE 7. Confidence intervals, mean and standard deviations for the time-invariant parameters, computed based on 1000 bootstrap samples using residual bootstrap approach for *Minnesota*.

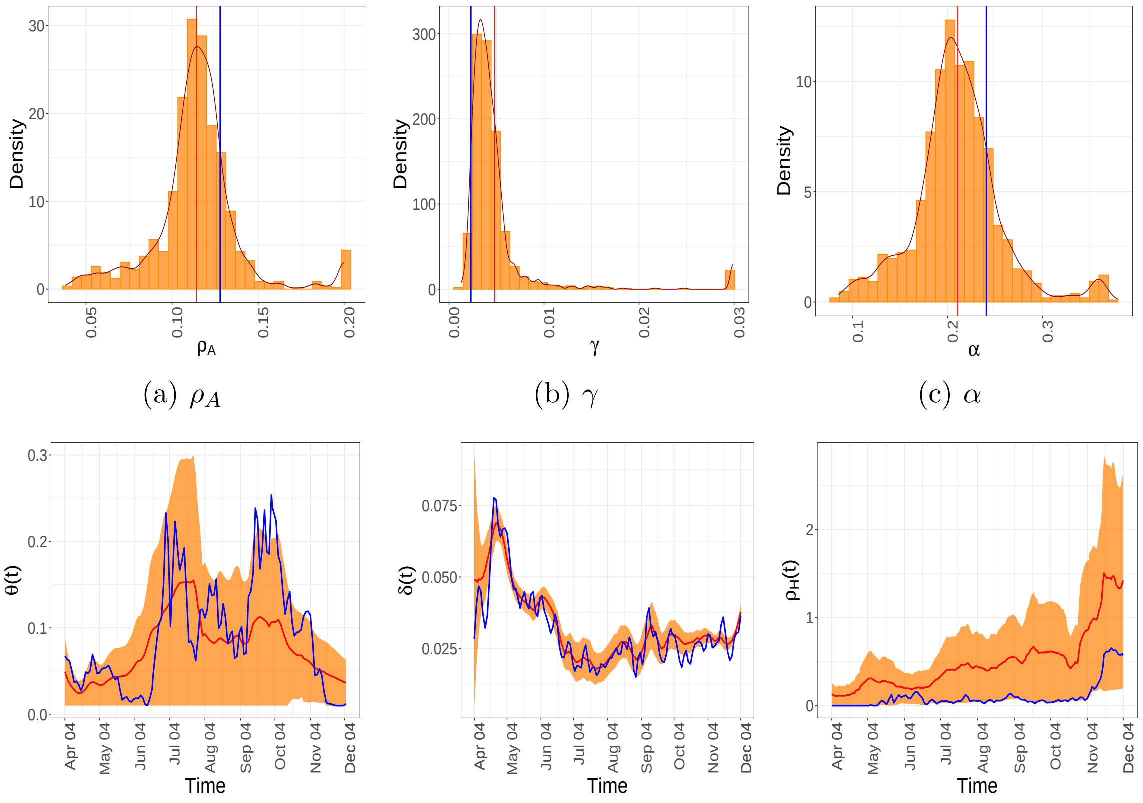


(a) $\rho_A$ (b) $\gamma$ (c) $\alpha$

(d) $\theta(t)$ (e) $\delta(t)$ (f) $\rho_H(t)$

FIGURE 18. Estimates and residual bootstrap based confidence intervals for time invariant and time-varying parameters for the state of *Minnesota*. The estimate from the data is in blue. The 95% confidence band is in yellow and the mean of the bootstrap estimates are presented in red.

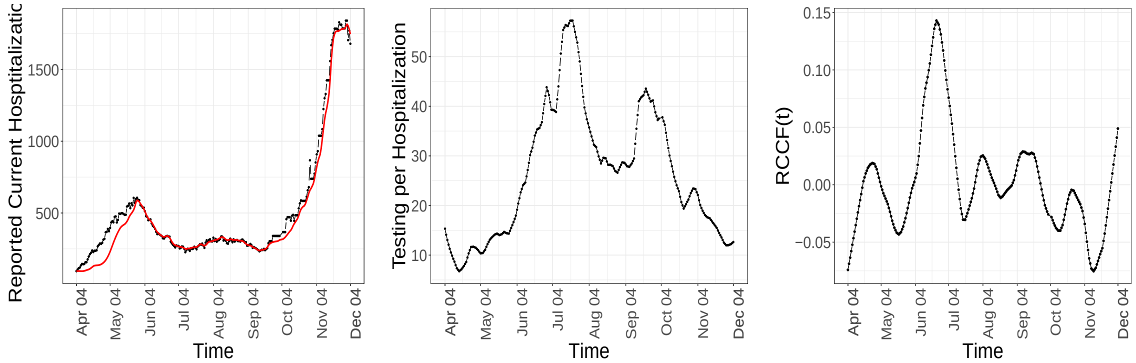


(a) Daily Hospitalisations, observed (–), fitted (–)

(b) $\Delta T_t/H_t$,

(c) RCCF

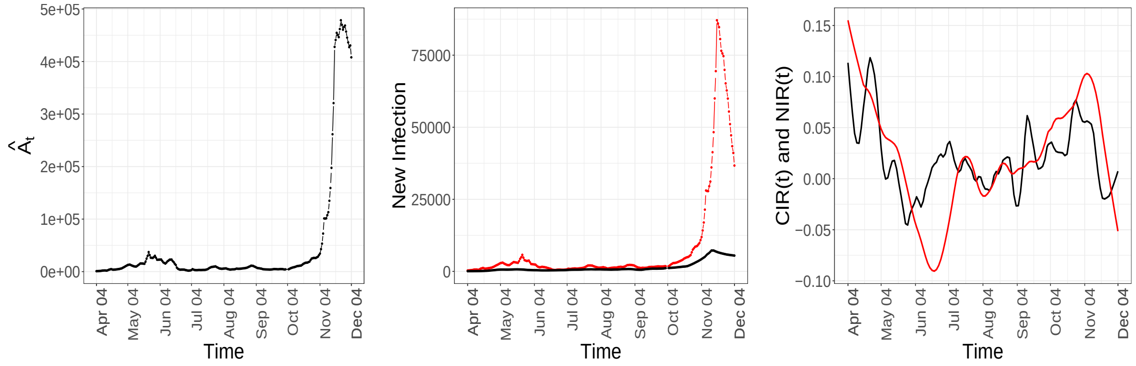


(d) $\hat{A}_t$

(e) Daily New Infections, observed (–), fitted (–)

(f) CIR (–) and NIR (–)

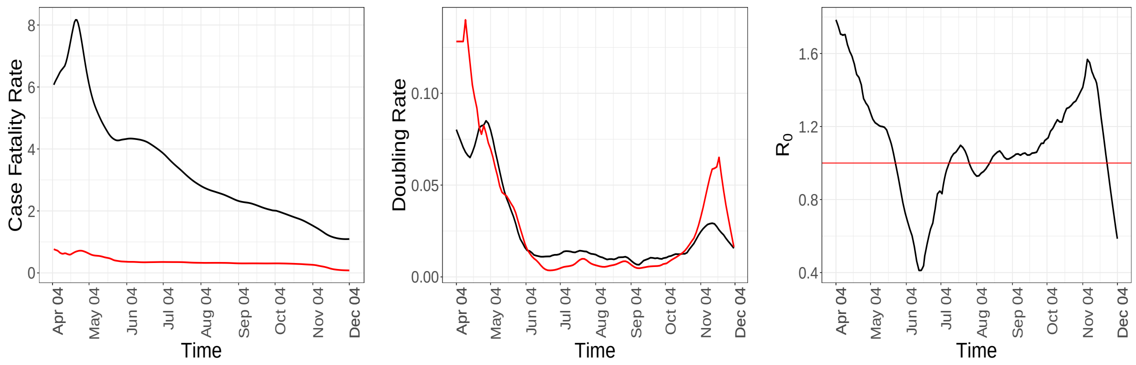


(g) Fatality Rate, observed (–), actual (–)

(h) Doubling rate, $C_t$ (–) and $CNI$ (–)

(i) $R_0$ analogue

FIGURE 19. Temporal patterns of some components and epidemiological markers for *Minnesota*.

### Nebraska

| | Estimate | 95% Confidence Interval | Mean | s.d. |
|---|---|---|---|---|
| $\gamma$ | 0.0011 | [0.0007, 0.0023] | 0.0013 | 0.0004 |
| $\rho_A$ | 0.0200 | [0.0160, 0.0300] | 0.0215 | 0.0003 |
| $\alpha$ | 0.0557 | [0.0399, 0.0969] | 0.0594 | 0.0138 |

TABLE 8. Confidence intervals, mean and standard deviations for the time-invariant parameters, computed based on 1000 bootstrap samples using residual bootstrap approach for *Nebraska*.

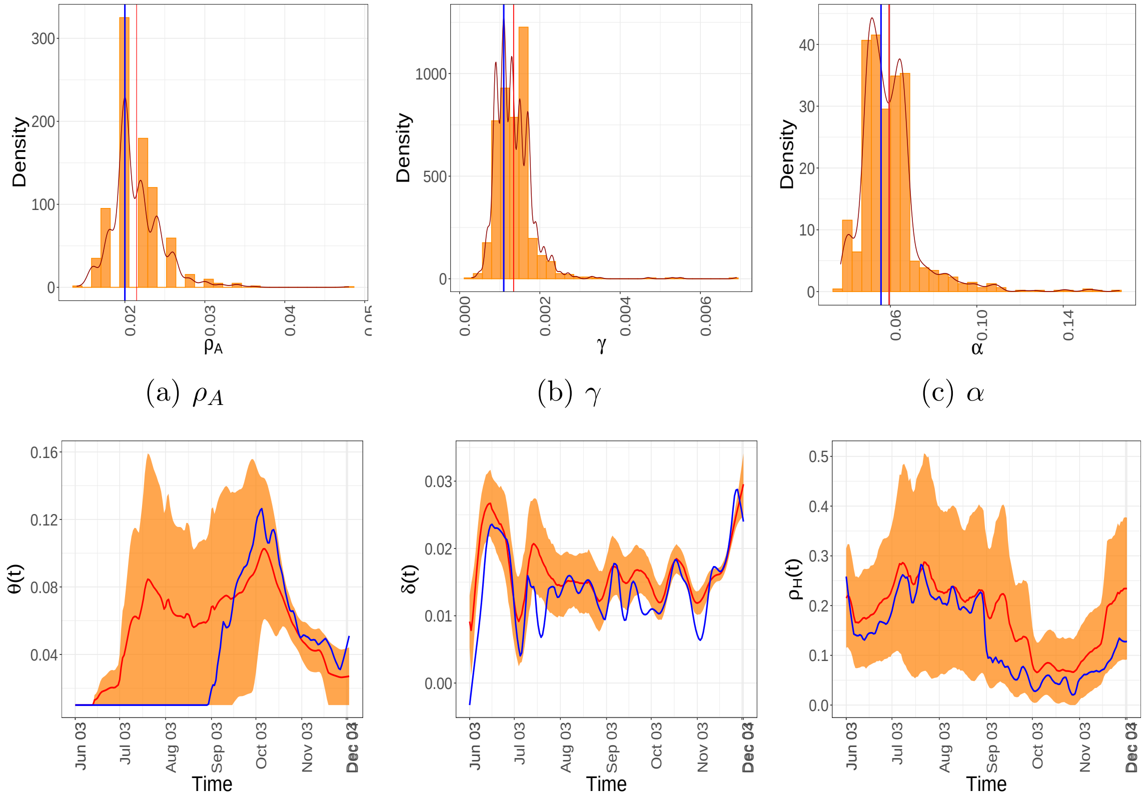


(a) $\rho_A$ (b) $\gamma$ (c) $\alpha$

(d) $\theta(t)$ (e) $\delta(t)$ (f) $\rho_H(t)$

FIGURE 20. Estimates and residual bootstrap based confidence intervals for time invariant and time-varying parameters for the state of *Nebraska*. The estimate from the data is in blue. The 95% confidence band is in yellow and the mean of the bootstrap estimates are presented in red.

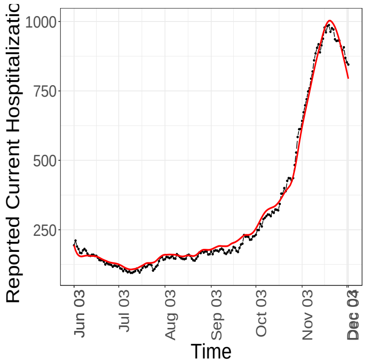


(a) Daily Hospitalisations, observed (—), fitted (—)

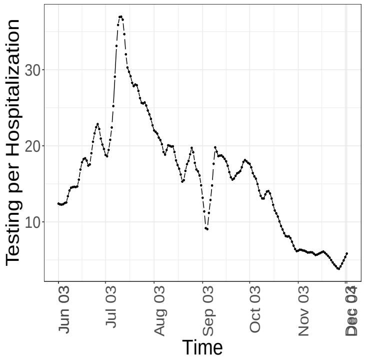


(b) $\Delta T_t/H_t$,

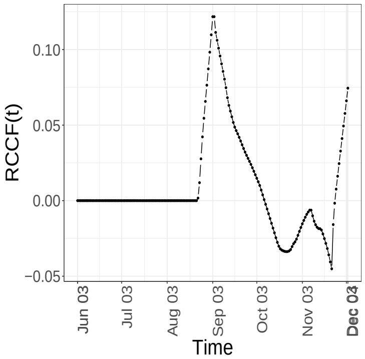


(c) RCCF

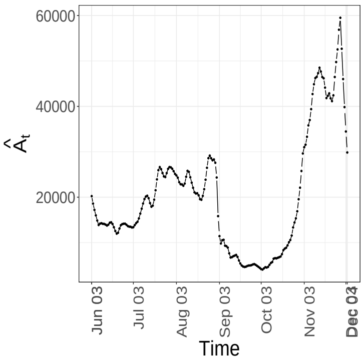


(d) $\hat{A}_t$

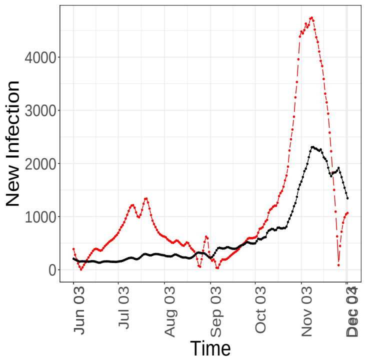


(e) Daily New Infections, observed (—), fitted (—)

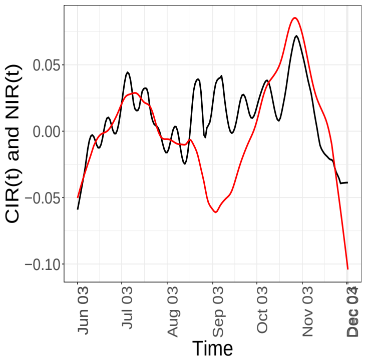


(f) CIR (—) and NIR (—)

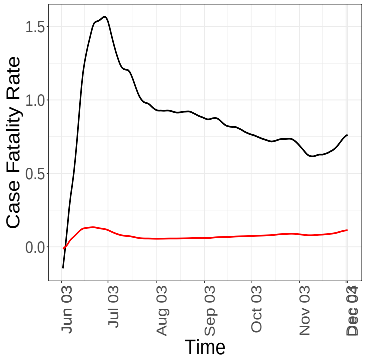


(g) Fatality Rate, observed (—), actual (—)

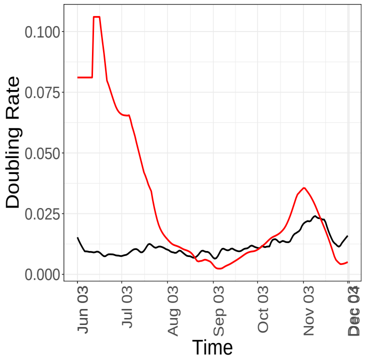


(h) Doubling rate, $C_t$ (—) and $CNI$ (—)

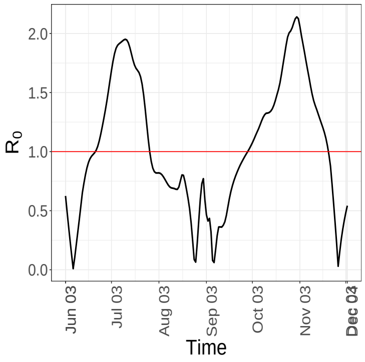


(i) $R_0$ analogue

FIGURE 21. Temporal patterns of some components and epidemiological markers for *Nebraska*.

### Ohio

| | Estimate | 95% Confidence Interval | Mean | s.d. |
|---|---|---|---|---|
| $\gamma$ | 0.0023 | [0.0023, 0.0037] | 0.0030 | 0.0003 |
| $\rho_A$ | 0.0480 | [0.0420, 0.0480] | 0.0458 | 0.0016 |
| $\alpha$ | 0.1027 | [0.0875, 0.1277] | 0.1050 | 0.0101 |

TABLE 9. Confidence intervals, mean and standard deviations for the time-invariant parameters, computed based on 1000 bootstrap samples using residual bootstrap approach for *Ohio*.

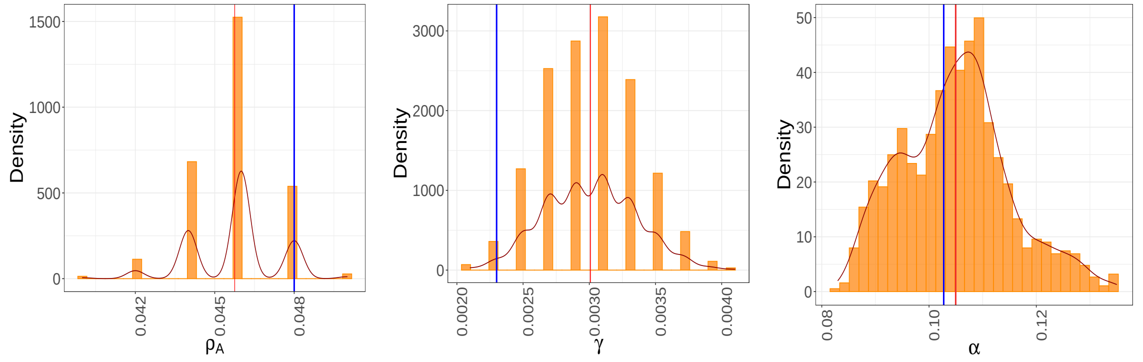


(a) $\rho_A$ (b) $\gamma$ (c) $\alpha$

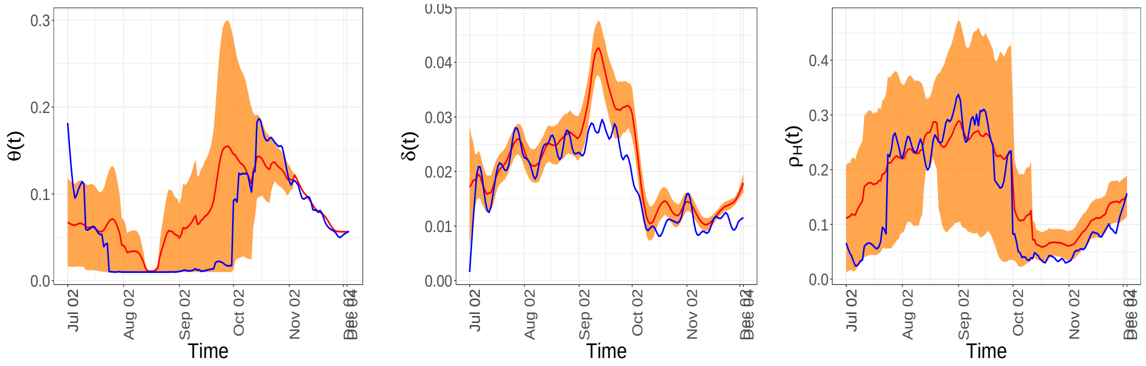


(d) $\theta(t)$ (e) $\delta(t)$ (f) $\rho_H(t)$

FIGURE 22. Estimates and residual bootstrap based confidence intervals for time invariant and time-varying parameters for the state of *Ohio*. The estimate from the data is in blue. The 95% confidence band is in yellow and the mean of the bootstrap estimates are presented in red.

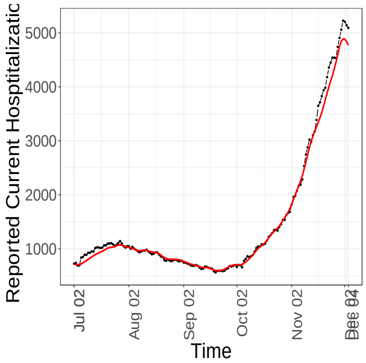


(a) Daily Hospitalisations, observed (—), fitted (—)

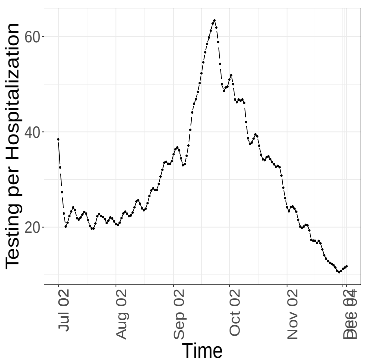


(b) $\Delta T_t/H_t$,

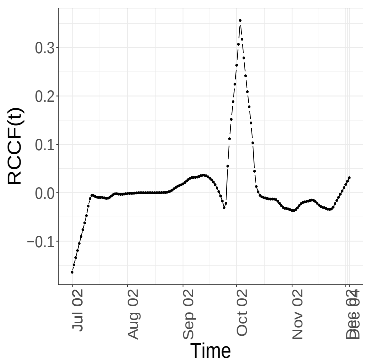


(c) RCCF

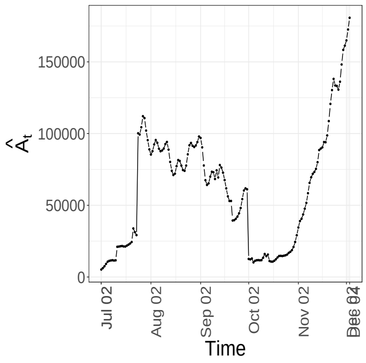


(d) $\hat{A}_t$

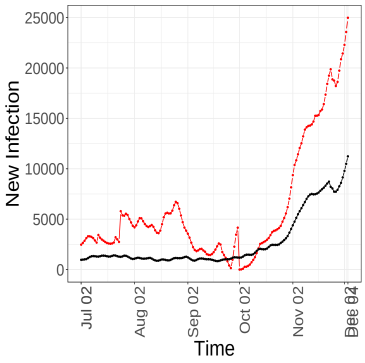


(e) Daily New Infections, observed (—), fitted (—)

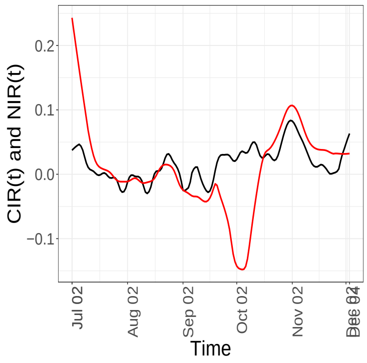


(f) CIR (—) and NIR (—)

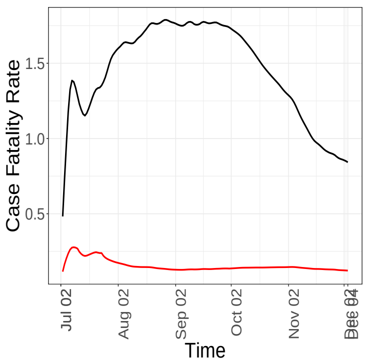


(g) Fatality Rate, observed (—), actual (—)

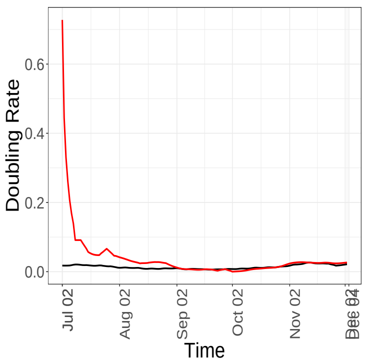


(h) Doubling rate, $C_t$ (—) and $CNI$ (—)

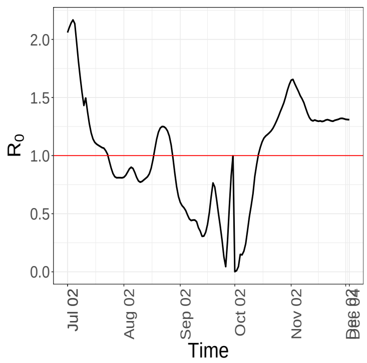


(i) $R_0$ analogue

FIGURE 23. Temporal patterns of some components and epidemiological markers for *Ohio*.

### Oklahoma

| | Estimate | 95% Confidence Interval | Mean | s.d. |
|---|---|---|---|---|
| $\gamma$ | 0037 | [0.0027, 0.0077] | 0.0053 | 0.0012 |
| $\rho_A$ | 0.0840 | [0.0660, 0.0840] | 0.0766 | 0.0044 |
| $\alpha$ | 0.1372 | [0.1477, 0.2089] | 0.1787 | 0.0138 |

TABLE 10. Confidence intervals, mean and standard deviations for the time-invariant parameters, computed based on 1000 bootstrap samples using residual bootstrap approach for *Oklahoma*.

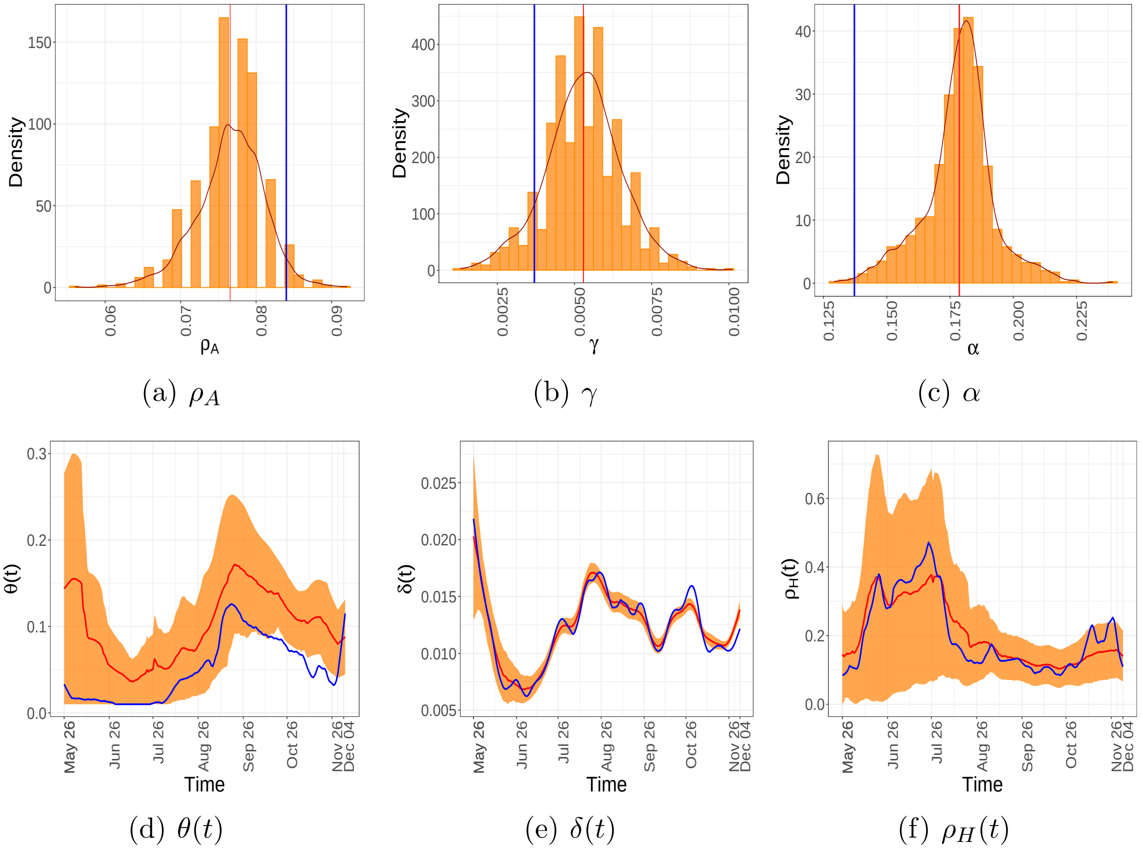


(a) $\rho_A$ (b) $\gamma$ (c) $\alpha$

(d) $\theta(t)$ (e) $\delta(t)$ (f) $\rho_H(t)$

FIGURE 24. Estimates and residual bootstrap based confidence intervals for time invariant and time-varying parameters for the state of *Oklahoma*. The estimate from the data is in blue. The 95% confidence band is in yellow and the mean of the bootstrap estimates are presented in red.

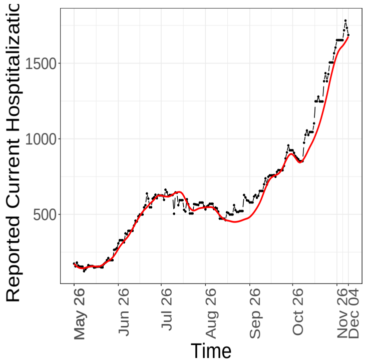


(a) Daily Hospitalisations, observed (–), fitted (–)

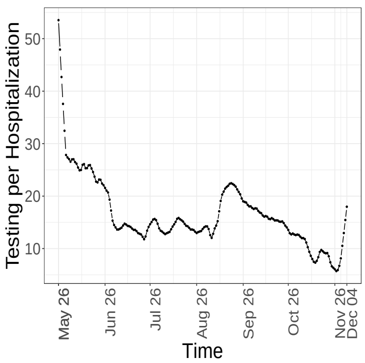


(b) $\Delta T_t/H_t$,

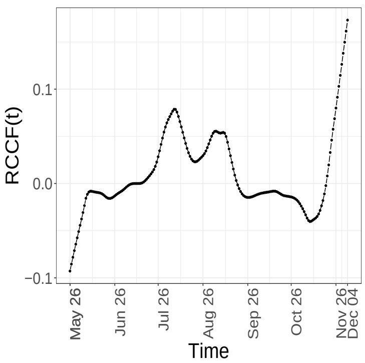


(c) RCCF

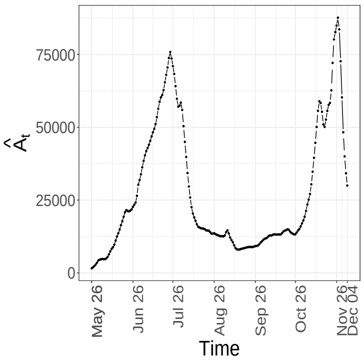


(d) $\hat{A}_t$

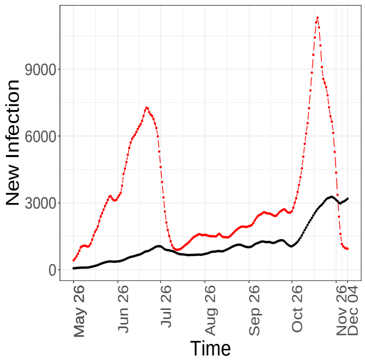


(e) Daily New Infections, observed (–), fitted (–)

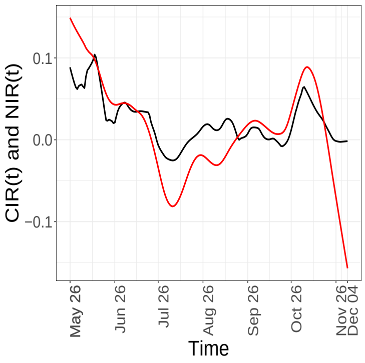


(f) CIR (–) and NIR (–)

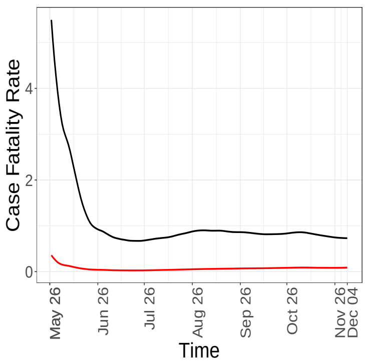


(g) Fatality Rate, observed (–), actual (–)

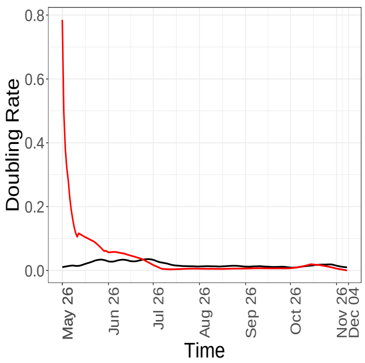


(h) Doubling rate, $C_t$ (–) and $CNI$ (–)

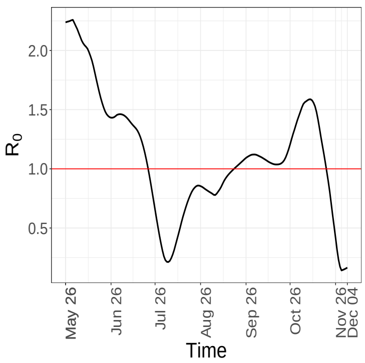


(i) $R_0$ analogue

FIGURE 25. Temporal patterns of some components and epidemiological markers for *Oklahoma*.

### Pennsylvania

| | Estimate | 95% Confidence Interval | Mean | s.d. |
|---|---|---|---|---|
| $\gamma$ | 0.0013 | [0.0013, 0.0023] | 0.0017 | 0.0003 |
| $\rho_A$ | 0.0260 | [0.0260, 0.0280] | 0.0274 | 0.0010 |
| $\alpha$ | 0.0818 | [0.0826, 0.1367] | 0.1085 | 0.0138 |

TABLE 11. Confidence intervals, mean and standard deviations for the time-invariant parameters, computed based on 1000 bootstrap samples using residual bootstrap approach for *Pennsylvania*.

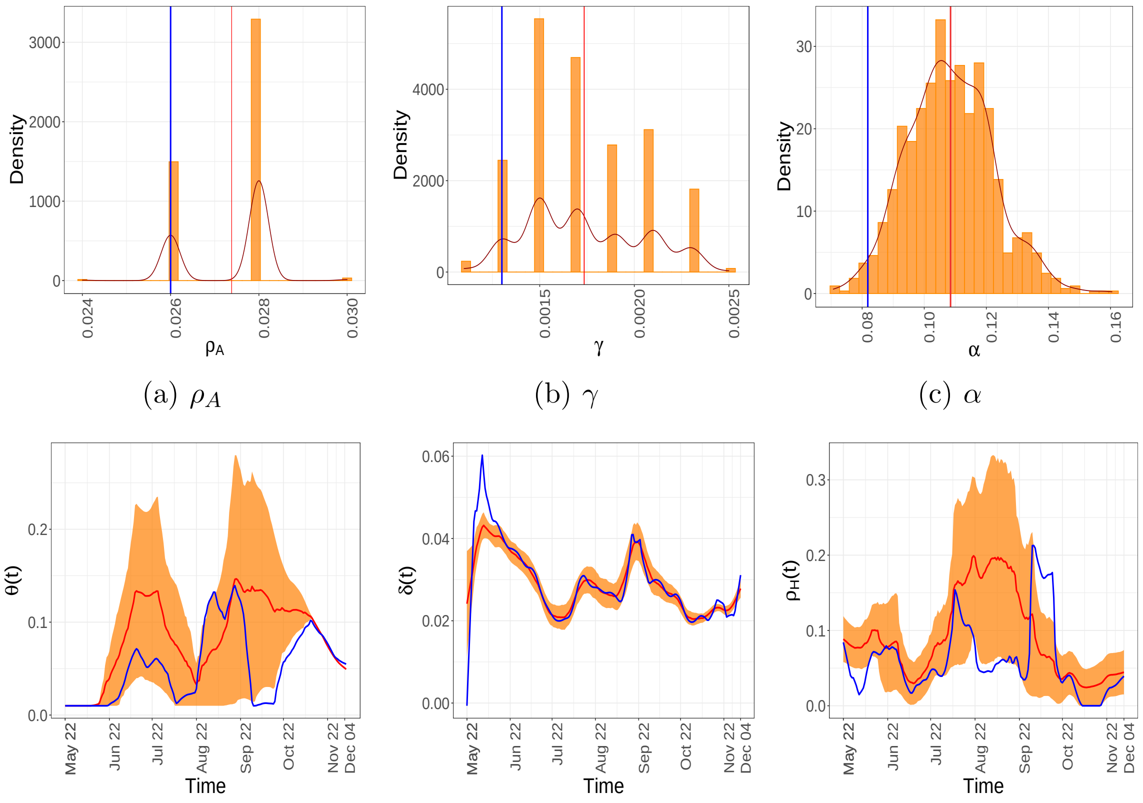


(a) $\rho_A$ (b) $\gamma$ (c) $\alpha$

(d) $\theta(t)$ (e) $\delta(t)$ (f) $\rho_H(t)$

FIGURE 26. Estimates and residual bootstrap based confidence intervals for time invariant and time-varying parameters for the state of *Pennsylvania*. The estimate from the data is in blue. The 95% confidence band is in yellow and the mean of the bootstrap estimates are presented in red.

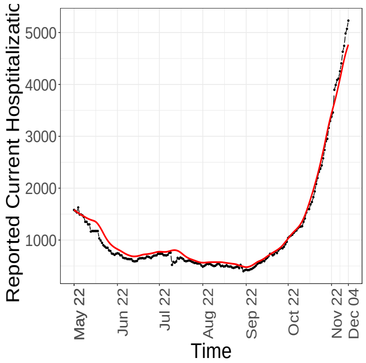


(a) Daily Hospitalisations, observed (–), fitted (–)

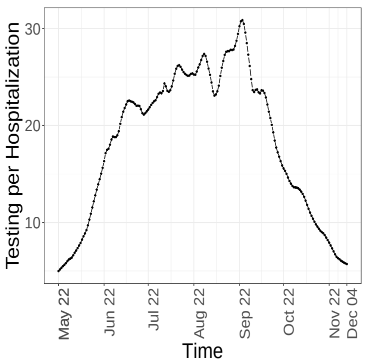


(b) $\Delta T_t/H_t$,

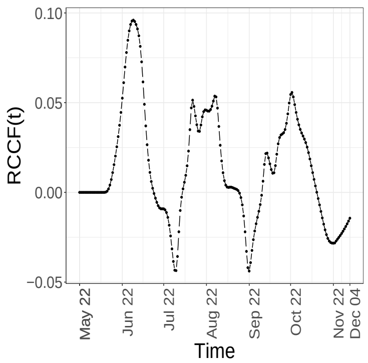


(c) RCCF

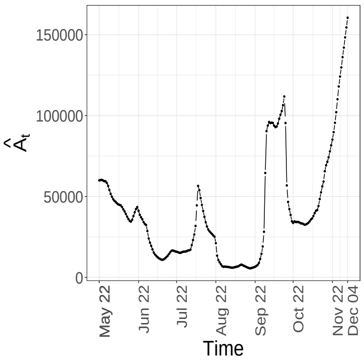


(d) $\hat{A}_t$

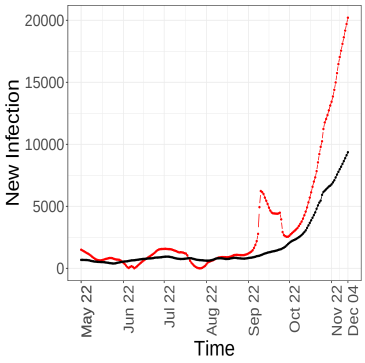


(e) Daily New Infections, observed (–), fitted (–)

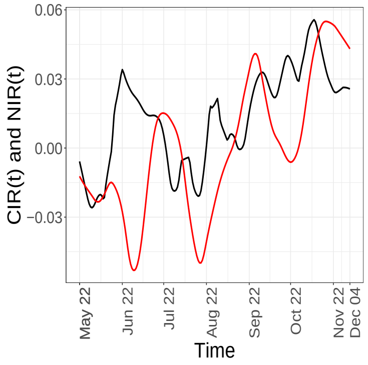


(f) CIR (–) and NIR (–)

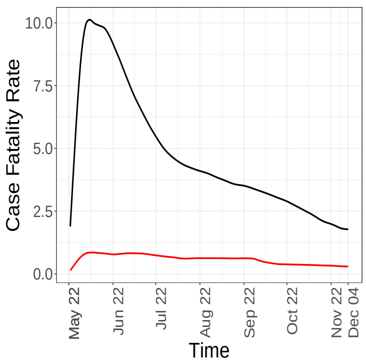


(g) Fatality Rate, observed (–), actual (–)

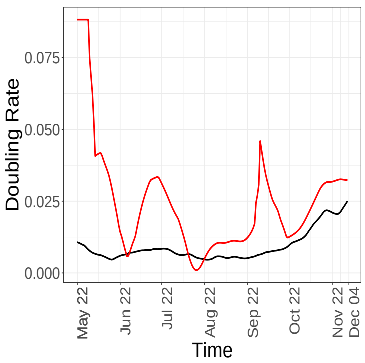


(h) Doubling rate, $C_t$ (–) and $CNI$ (–)

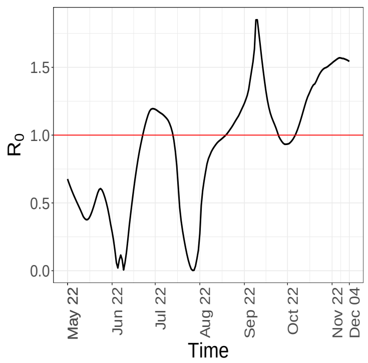


(i) $R_0$ analogue

FIGURE 27. Temporal patterns of some components and epidemiological markers for *Pennsylvania*.

### South Dakota

| | Estimate | 95% Confidence Interval | Mean | s.d. |
|---|---|---|---|---|
| $\gamma$ | 0.0021 | [0.0011, 0.0035] | 0.0022 | 0.0006 |
| $\rho_A$ | 0.0580 | [0.0580, 0.0740] | 0.0664 | 0.0041 |
| $\alpha$ | 0.0561 | [0.0668, 0.1053] | 0.0863 | 0.0101 |

TABLE 12. Confidence intervals, mean and standard deviations for the time-invariant parameters, computed based on 1000 bootstrap samples using residual bootstrap approach for *South Dakota*.

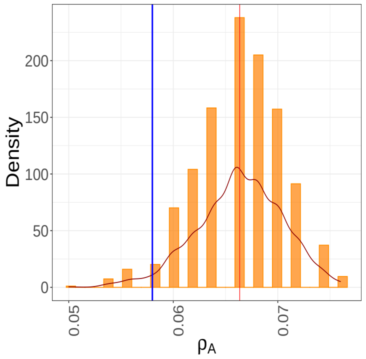


(a) $\rho_A$

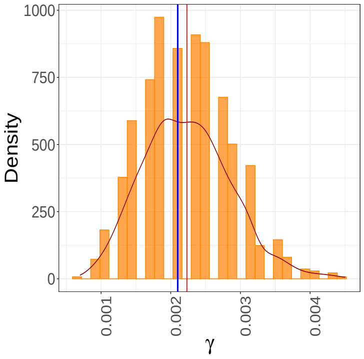


(b) $\gamma$

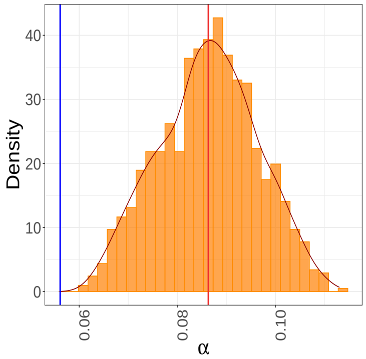


(c) $\alpha$

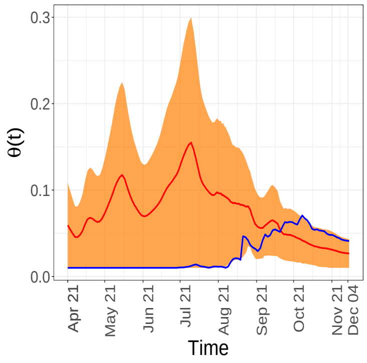


(d) $\theta(t)$

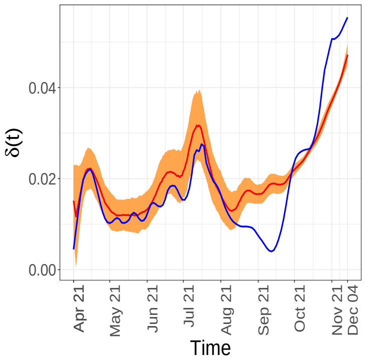


(e) $\delta(t)$

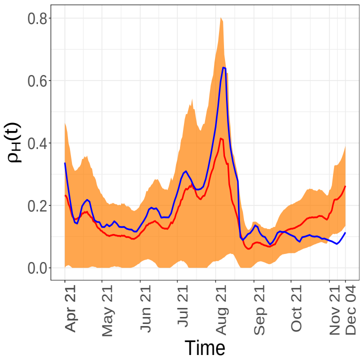


(f) $\rho_H(t)$

FIGURE 28. Estimates and residual bootstrap based confidence intervals for time invariant and time-varying parameters for the state of *South Dakota*. The estimate from the data is in blue. The 95% confidence band is in yellow and the mean of the bootstrap estimates are presented in red.

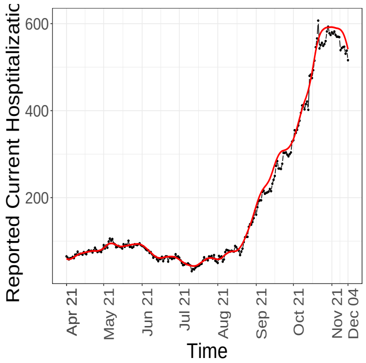


(a) Daily Hospitalisations, observed (–), fitted (–)

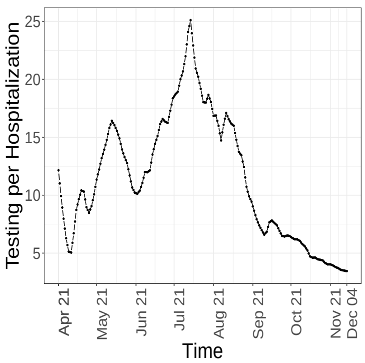


(b) $\Delta T_t/H_t$,

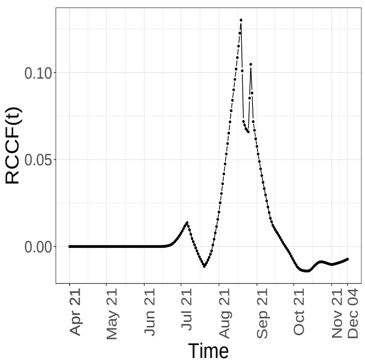


(c) RCCF

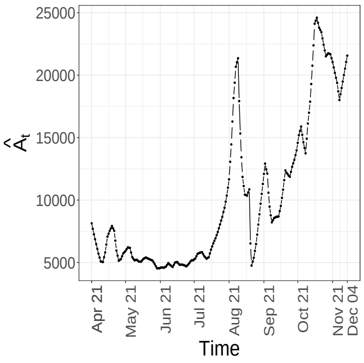


(d) $\hat{A}_t$

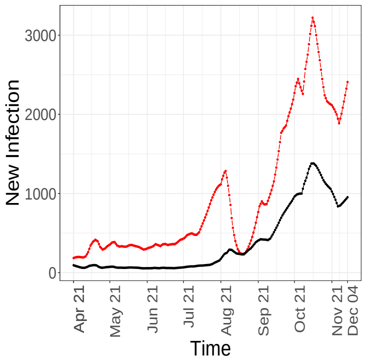


(e) Daily New Infections, observed (–), fitted (–)

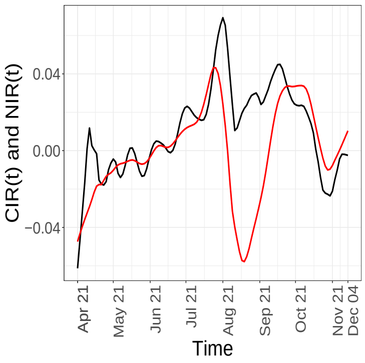


(f) CIR (–) and NIR (–)

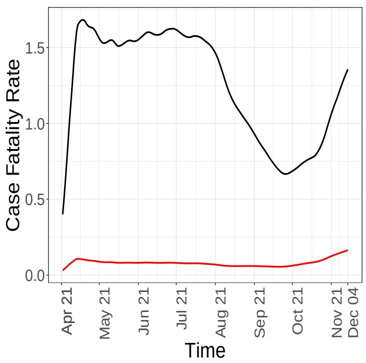


(g) Fatality Rate, observed (–), actual (–)

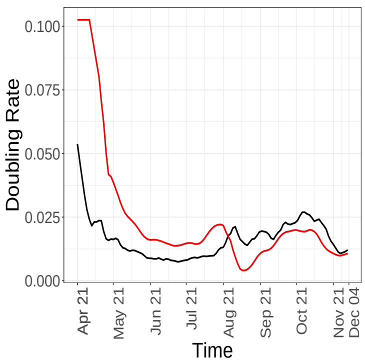


(h) Doubling rate, $C_t$ (–) and $CNI$ (–)

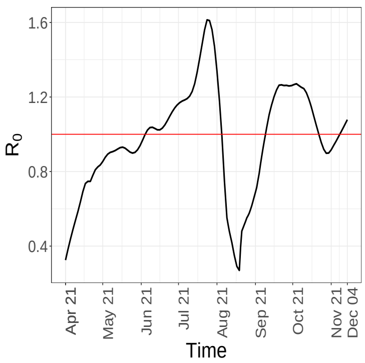


(i) $R_0$ analogue

FIGURE 29. Temporal patterns of some components and epidemiological markers for *South Dakota*.

### Tennessee

| | Estimate | 95% Confidence Interval | Mean | s.d. |
|---|---|---|---|---|
| $\gamma$ | 0.0059 | [0.0059, 0.0075] | 0.0066 | 0.0004 |
| $\rho_A$ | 0.0640 | [0.0560, 0.0660] | 0.6081 | 0.0025 |
| $\alpha$ | 0.1907 | [0.1564, 0.1861] | 0.1710 | 0.0076 |

TABLE 13. Confidence intervals, mean and standard deviations for the time-invariant parameters, computed based on 1000 bootstrap samples using residual bootstrap approach for *Tennessee*.

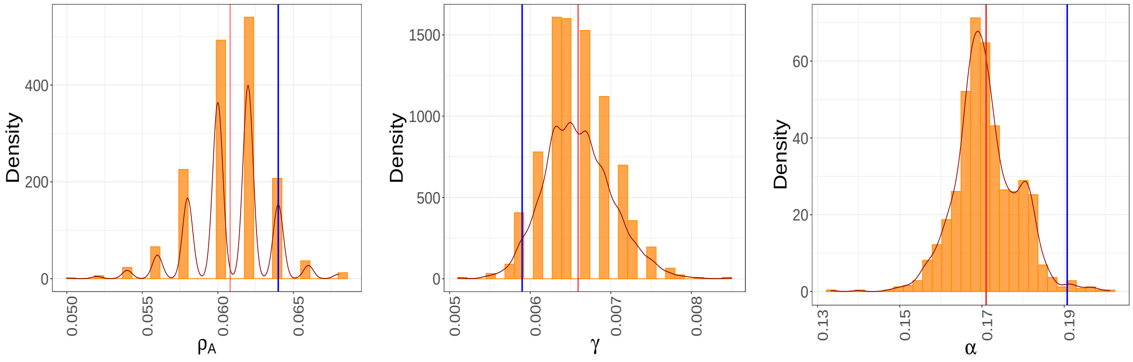


(a) $\rho_A$ (b) $\gamma$ (c) $\alpha$

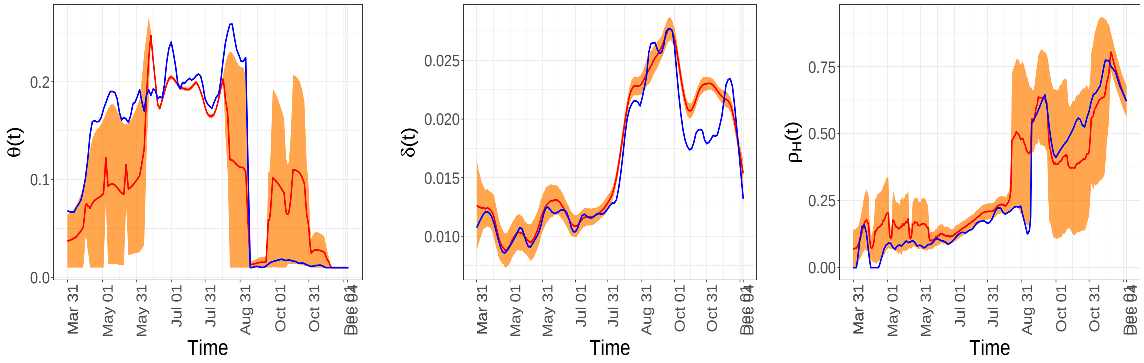


(d) $\theta(t)$ (e) $\delta(t)$ (f) $\rho_H(t)$

FIGURE 30. Estimates and residual bootstrap based confidence intervals for time invariant and time-varying parameters for the state of *Tennessee*. The estimate from the data is in blue. The 95% confidence band is in yellow and the mean of the bootstrap estimates are presented in red.

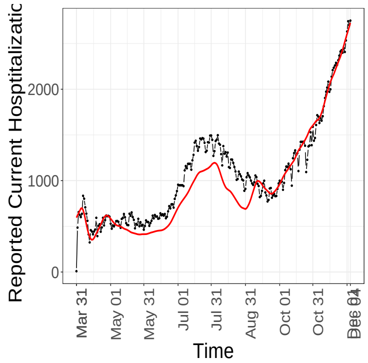


(a) Daily Hospitalisations, observed (−), fitted (−)

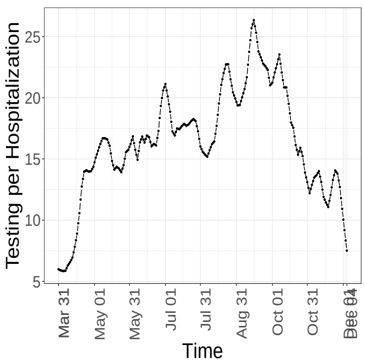


(b) $\Delta T_t/H_t$,

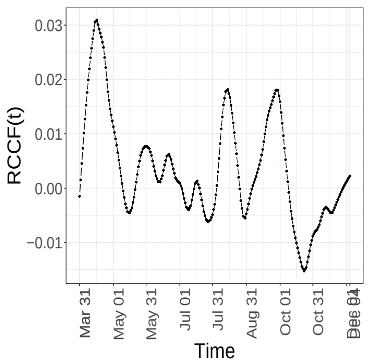


(c) RCCF

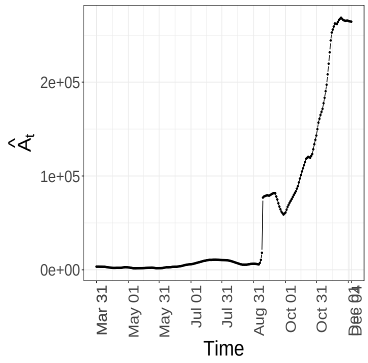


(d) $\hat{A}_t$

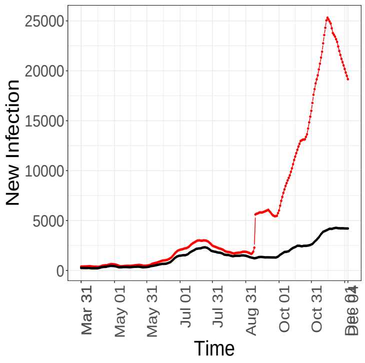


(e) Daily New Infections, observed (−), fitted (−)

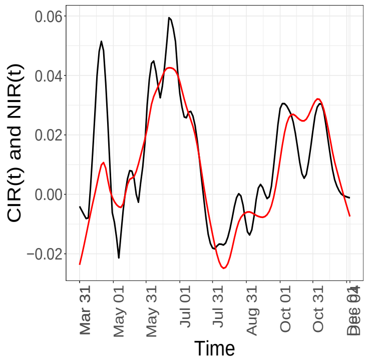


(f) CIR (−) and NIR (−)

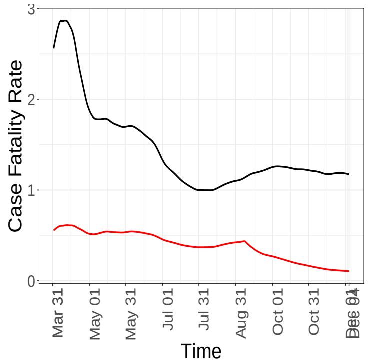


(g) Fatality Rate, observed (−), actual (−)

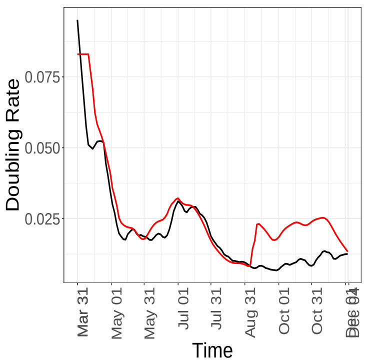


(h) Doubling rate, $C_t$ (−) and $CNI$ (−)

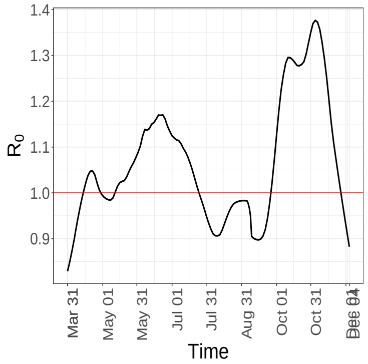


(i) $R_0$ analogue

FIGURE 31. Temporal patterns of some components and epidemiological markers for *Tennessee*.

### Texas

| | Estimate | 95% Confidence Interval | Mean | s.d. |
|---|---|---|---|---|
| $\gamma$ | 0.0019 | [0.0019, 0.0027] | 0.0023 | 0.0003 |
| $\rho_A$ | 0.0360 | [0.0320, 0.0360] | 0.0348 | 0.0013 |
| $\alpha$ | 0.0986 | [0.1021, 0.1583] | 0.1325 | 0.0142 |

TABLE 14. Confidence intervals, mean and standard deviations for the time-invariant parameters, computed based on 1000 bootstrap samples using residual bootstrap approach for *Texas*.

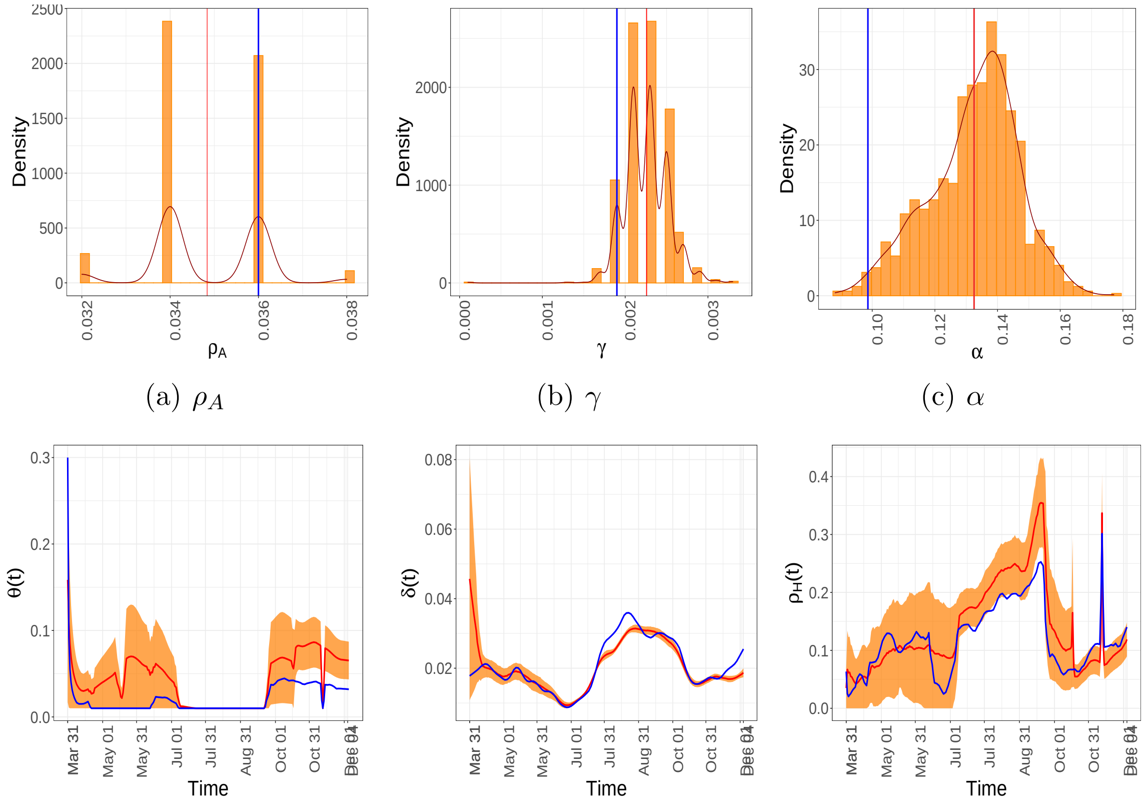


(a) $\rho_A$ (b) $\gamma$ (c) $\alpha$

(d) $\theta(t)$ (e) $\delta(t)$ (f) $\rho_H(t)$

FIGURE 32. Estimates and residual bootstrap based confidence intervals for time invariant and time-varying parameters for the state of *Texas*. The estimate from the data is in blue. The 95% confidence band is in yellow and the mean of the bootstrap estimates are presented in red.

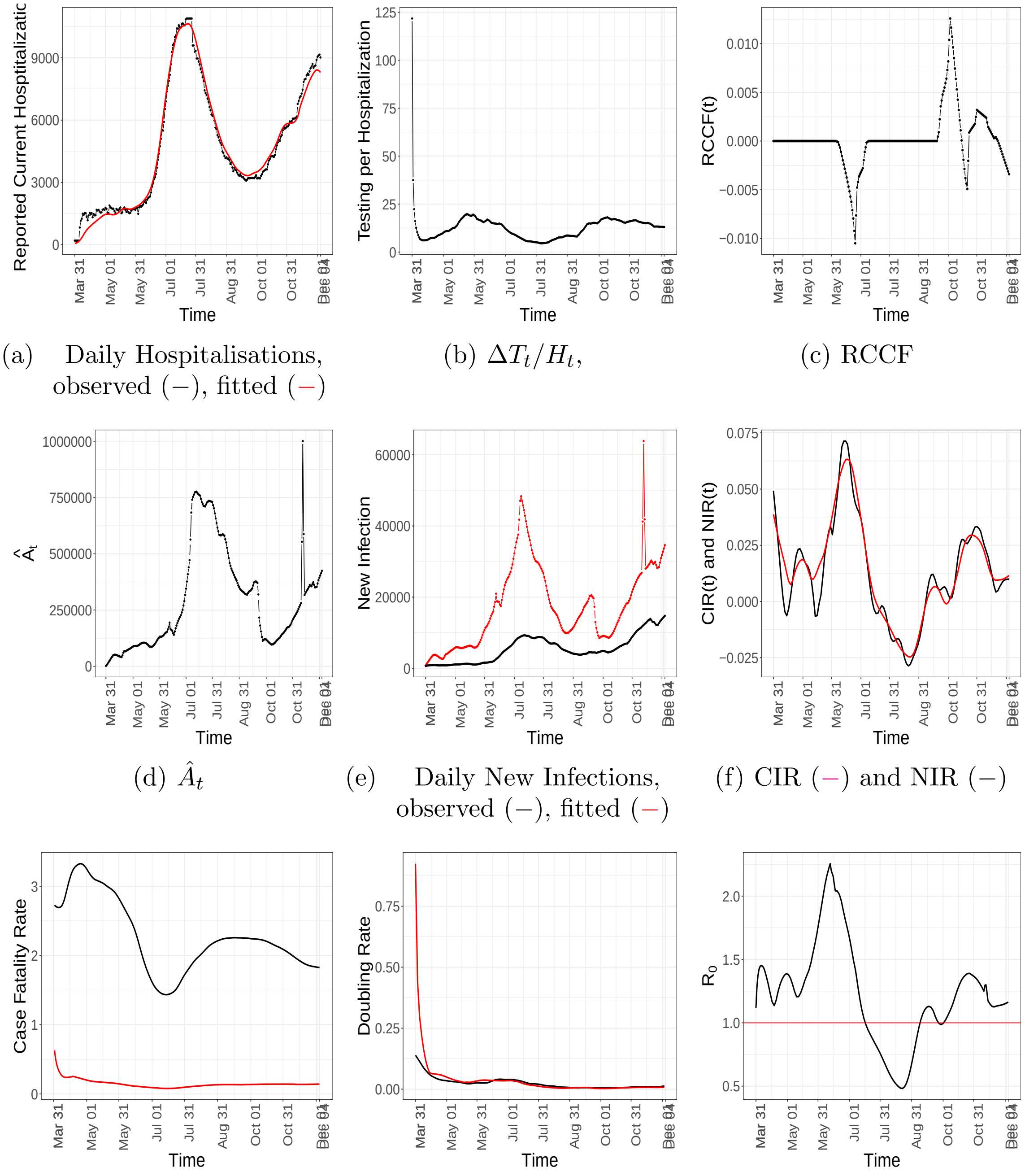


(a) Daily Hospitalisations, observed (–), fitted (–)

(b) $\Delta T_t / H_t$,

(c) RCCF

(d) $\hat{A}_t$

(e) Daily New Infections, observed (–), fitted (–)

(f) CIR (–) and NIR (–)

(g) Fatality Rate, observed (–), actual (–)

(h) Doubling rate, $C_t$ (–) and $CNI$ (–)

(i) $R_0$ analogue

FIGURE 33. Temporal patterns of some components and epidemiological markers for *Texas*.

### Wisconsin

| | Estimate | 95% Confidence Interval | Mean | s.d. |
|---|---|---|---|---|
| $\gamma$ | 0.0017 | [0.0015, 0.0033] | 0.0024 | 0.0040 |
| $\rho_A$ | 0.0680 | [0.0600, 0.0700] | 0.0646 | 0.0027 |
| $\alpha$ | 0.0926 | [0.0761, 0.1207] | 0.1000 | 0.0121 |

TABLE 15. Confidence intervals, mean and standard deviations for the time-invariant parameters, computed based on 1000 bootstrap samples using residual bootstrap approach for *Wisconsin*.

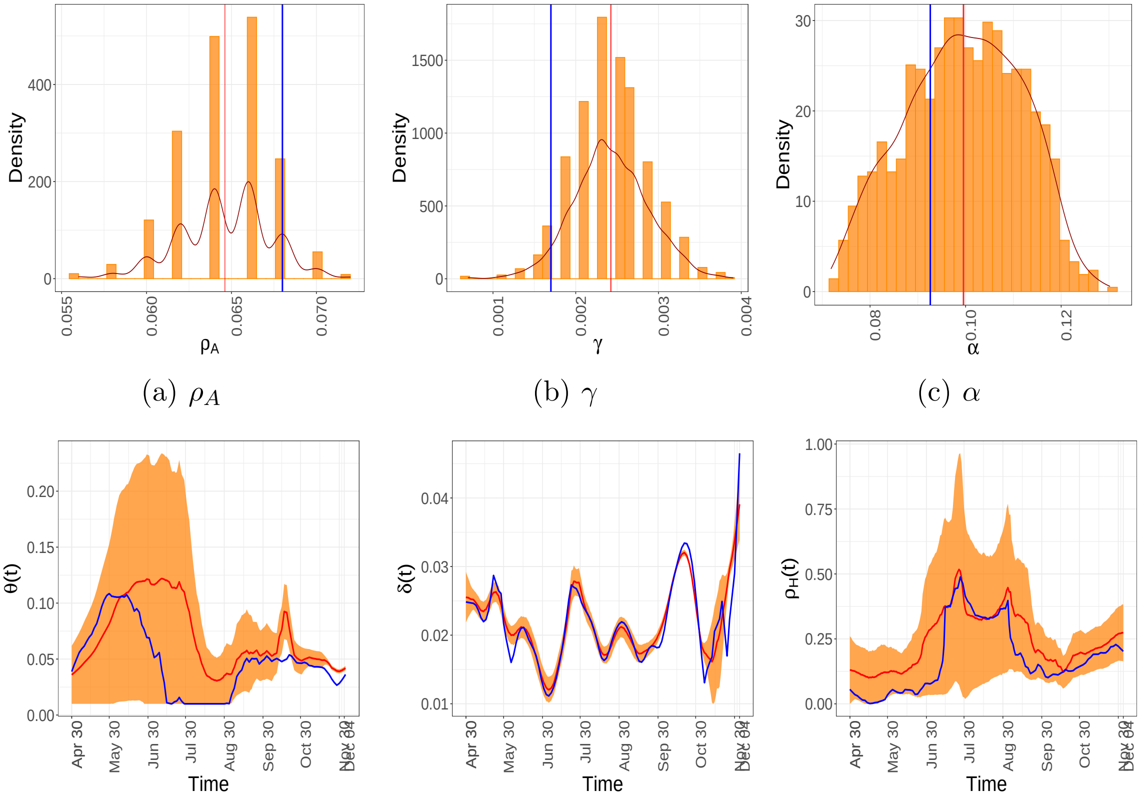


(a) $\rho_A$ (b) $\gamma$ (c) $\alpha$

(d) $\theta(t)$ (e) $\delta_t$ (f) $\rho_H(t)$

FIGURE 34. Estimates and residual bootstrap based confidence intervals for time invariant and time-varying parameters for the state of *Wisconsin*. The estimate from the data is in blue. The 95% confidence band is in yellow and the mean of the bootstrap estimates are presented in red.

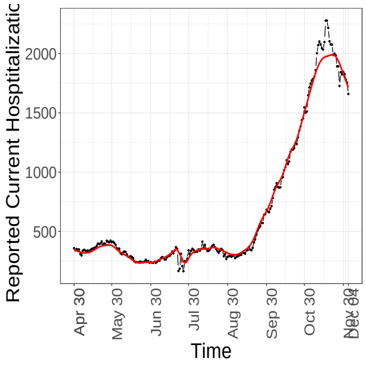


(a) Daily Hospitalisations, observed (−), fitted (−)

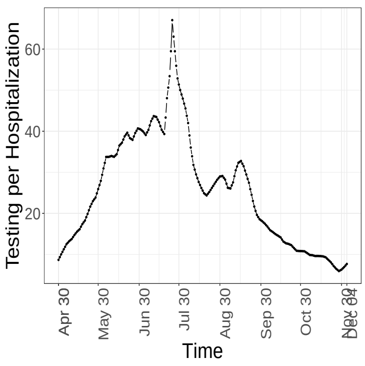


(b) $\Delta T_t/H_t$,

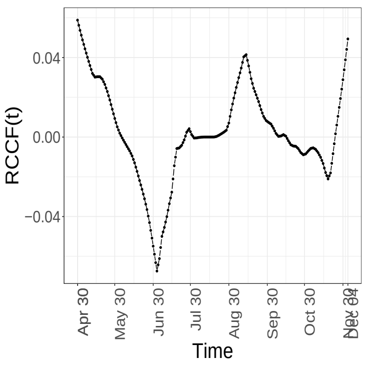


(c) RCCF

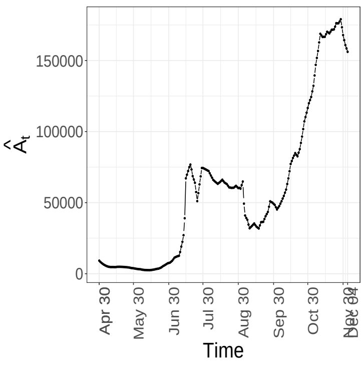


(d) $\hat{A}_t$

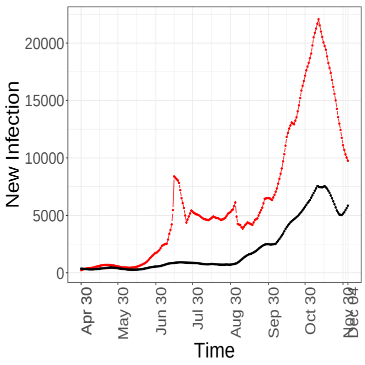


(e) Daily New Infections, observed (−), fitted (−)

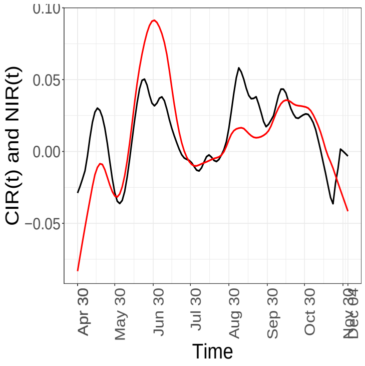


(f) CIR (−) and NIR (−)

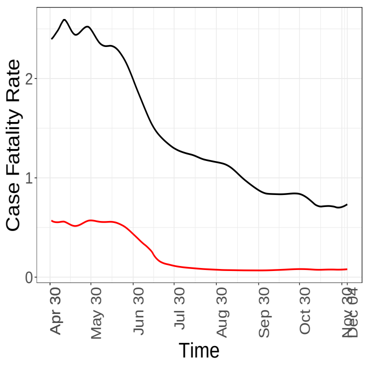


(g) Fatality Rate, observed (−), actual (−)

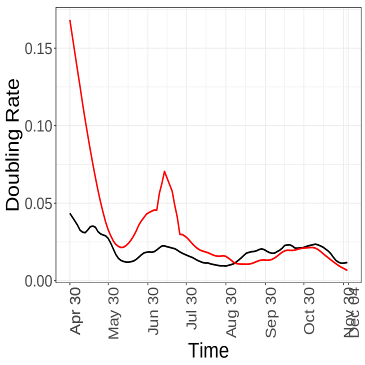


(h) Doubling rate, $C_t$ (−) and $CNI$ (−)

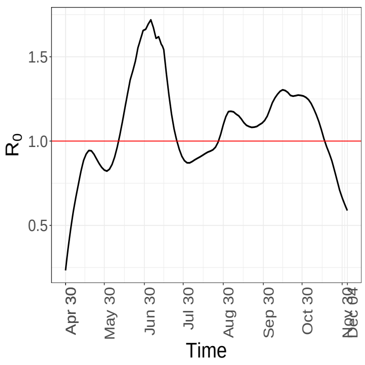


(i) $R_0$ analogue

Figure 35. Temporal patterns of some components and epidemiological markers for *Wisconsin*.
